# Global DNA methylomes reveal oncogenic-associated 5-hydroxylmethylated cytosine (5hmC) signatures in the cell-free DNA of cancer patients

**DOI:** 10.1101/2025.01.09.25320283

**Authors:** Gabriel E. Rech, Alyssa C. Lau, Rachel L. Goldfeder, Rahul Maurya, Alexey V. Danilov, Chia-Lin Wei

## Abstract

Characterization of tumor epigenetic aberrations is integral to understanding the mechanisms of tumorigenesis and provide diagnostic, prognostic, and predictive information of high clinical relevance. Among the different tumor-associated epigenetic signatures, 5 methyl-cytosine (5mC) and 5-hydroxymethylcytosine (5hmC) are the two most well-characterized DNA methylation alterations linked to cancer pathogenesis. 5hmC has a tissue-specific distribution and its abundance is subjected to changes in tumor DNA, making it a promising biomarker. Detecting tumor-related DNA methylation alterations in tissues is highly invasive, while the analysis of the cell-free DNA (cfDNA) is poised to supplement, if not replace, surgical biopsies. Despite many studies attempted to identify new epigenetic targets for liquid biopsy assays, little is known about the regulatory roles of 5hmC, its impacts on the molecular phenotypes in tumors. Most importantly, whether the oncogenic-associated 5hmC signatures found in tumor tissues can be recapitulated in patients’ cfDNA. In this study, we performed the unbiased and simultaneous detection of 5mC and 5hmC whole-genome DNA modifications at base-resolution from two distinct cancer cohorts, from patients with bladder cancer or B-Cell lymphoma, their corresponding normal tissues, and cfDNAs from plasma. We analyzed tissue-specific methylation patters and searched for signatures in gene coding and regulatory regions linked to cancerous states. We then looked for methylation signatures in patients’ cfDNA to determine if they were consistent with the tumor-specific patterns. We determined the functional significance of 5hmC in tissue specific transcription and uncovered hundreds of tumor-associated 5hmC signatures. These tumor-associated 5hmC changes, particularly in genes and enhancers, were functionally significant in tumorigenesis pathways and correlated with tumor specific gene expression. To investigate if cfDNA is a faithful surrogate for tumor-associated 5hmC, we devised a targeted capture strategy to examine the alterations of 5hmC in cfDNA from patients with bladder cancer and lymphoma with sufficient sensitivity and specificity and confirmed that they recapitulated the patterns we observed in tumor tissues. Our results provide analytic validation of 5hmC as a cancer-specific biomarker. The methods described here for systematic characterization of 5hmC at functional elements open new avenues to discover epigenetic markers for non-invasive diagnosis, monitoring, and stratifying cancer.

## Introduction

Aberrant DNA methylation is one of the most common hallmarks of human cancers [1] and it contributes to tumorigenesis across many different cancer types [2, 3]. Measuring the alterations of DNA methylation patterns in tumor tissues has been shown to provide valuable insight into their molecular abnormalities and therapeutic responses [4]. Such tumor-associated epigenetic signatures can serve as effective molecular marks to distinguish cancer from normal cells [1], making them a valuable tool for cancer diagnosis and prognosis. Among the different types of cancer-associated epigenetic signatures, 5 methyl-cytosine (5mC) and 5-hydroxymethylcytosine (5hmC) are the two most well-characterized types of DNA methylation linked to cancer pathogenesis [5, 6]. Both are found predominantly in the CpG context across the mammalian genomes. 5mC is highly abundant and associated with inactive chromatin states [7, 8]. Meanwhile, 5hmC is low abundant, often found in open chromatin regions and typically associated with active transcription [9]. Moreover, 5hmC has a tissue-specific distribution [10]. Recent studies have shown that 5hmC level and distribution are subject to changes in tumor DNA [11–15], and the aberration of 5hmC has been correlated with dysregulation of genes in oncogenic pathways [16]. In some solid tumors and *in vitro* cancer models, global loss of 5hmC was reported in tumor tissues when compared with their non-tumor counterparts [12–15]. Hence, the characterization of tumor-related 5hmC alterations in clinical samples offer potential utilities in cancer diagnosis, monitoring disease progression, and therapeutic responses [14, 17, 18].

In clinical settings, extracting tumor cells from cancer patients is highly invasive, particularly for solid tumors, whereas cell-free DNA (cfDNA), the DNA released from apoptotic and necrotic cells in the bloodstream or body fluids, can be isolated from the plasma. CfDNA carries the equivalent nucleotide context and modifications from its tissue origins, and therefore can provide snapshots into the genetic and epigenetic information of a patients’ tumor [19–21]. Known as liquid biopsy, analyzing cfDNA from cancer patients has offered promising potential as a minimal-invasive method for accessing patients’ tumor status and treatment responses [22]. Many types of cancer-associated genetic variations have been evaluated in cfDNA, including somatic mutations, copy number alterations (CNA) and breakpoints of structural variants [23–25]. Yet, the variable tumor fractions in cfDNA and its quantity in plasma present significant limitations for downstream analyses. By detecting only a handful of tumor-associated single nucleotide mutations or structural variants in cfDNA, it could be insufficient to achieve the required sensitivity and comprehensiveness to detect cancer cells and measure their malignant status. On the other hand, the cfDNA methylome serves as a comprehensive and dynamic readout of DNA activities throughout the body, offering a powerful tool for evaluating its changes associated with tumor origins and subtypes in cancer patients [26, 27]. Growing evidence has shown that cell-free methylomes exhibited tumor-specific changes associated with many solid cancer types [28–32] and provided insights into tumor heterogeneity and status, which is particularly useful for monitoring metastatic cancers [33]. Specifically, the tissue-specific distribution patterns of 5hmC make it a particularly valuable epigenetic marker in cfDNA analysis, especially in the context of cancer detection and classification [17, 34]. Hence, the analysis of cfDNA hydroxymethylomes, leveraging the tissue-specific nature, offers a promising approach for the sensitive and specific detection and classification of various cancer types [35].

To harness 5hmC’s potential in cancer detection and monitoring, many studies have aimed to identify cancer-specific 5hmC changes in cfDNA by comparing 5hmC profiles between large groups of patient and control cohorts. Such large population of sample sizes often lead to limitations in the scope of genomic analyses to only focus on a subset of the genomic regions [23, 28, 29, 34, 36] or selective sites enriched for high 5hmC levels [17, 33, 37, 38]. Little is known about the regulatory roles of 5hmC, its genome-wide changes and their impacts on the molecular phenotypes in tumors. Most importantly, whether the oncogenic-associated 5hmC signatures found in tumor tissues can be recapitulated in patients’ cfDNA. To address these knowledge gaps, we systematically profiled genome-wide 5hmc and 5mC methylomes simultaneously from bladder tissues and lymph node B-cells, together with their corresponding cancerous tissues: bladder tumors and non-Hodgkin lymphoma (NHL) tumors. We focused on these two malignancies because their tumor phenotypes have been found to associate with DNA methylation [17, 39, 40] and these cancers models are associated with distinct pathogenesis and clinical outcomes. From over 20 unbiased, genome-wide 5mC and 5hmC methylomes in bladder urothelial and lymph node B-cells DNA at single nucleotide resolution, we evaluated the tissue specificity of DNA methylation, uncovered the methylation signatures at both gene coding and regulatory regions associated with the oncogenic states, and observed their impact on differential transcription of genes in functional pathways affected by the malignancy. To examine if cfDNA can be a faithful surrogate for accessing these tumor specific methylation patterns, we assessed these differential methylation signatures in the patients’ cfDNAs and found them exhibit concordant changes when compared to cfDNAs from control individuals. Our data suggests that measuring 5hmC in cfDNA can act as a reliable readout for its variation in tumor biopsies and reveals potential epigenetic targets for developing tumor biomarkers. Taken together, our study offers solid supports for 5hmC as an effective signature to non-invasive diagnosis, monitoring, and stratification of cancer.

## Results

### Genome-wide, base-resolution DNA methylomes in malignant and paired healthy control urothelial and lymphoid tissues

To gain insights into the tissue-specificity of 5mC and 5hmC modifications, their changes associated with malignant states and potential values as biomarkers in plasma from cancer patients, we systematically surveyed 5mC and 5hmC methylomes at genome-wide, single-base resolution in two distinct cancer cohorts: bladder cancer patients and NHL patients. For bladder cancer, we analyzed tumor biopsies from five patients with stage pT3 high-grade muscle-invasive urothelial carcinoma (Tumor Bladder tissue samples) and their adjacent healthy urothelial tissues (Normal Bladder tissue samples). For NHL patients, we collected purified malignant B-cells from excisional lymph node biopsies (NHL Tumors, n=9). As control for NHL Tumors, we used healthy germinal center (GC) B-cells isolated from tonsils from healthy individuals (Normal B-Cells, n=5) [54]. For cfDNA survey, we collected plasma from bladder cancer patients at the same pT3 stage (n=8) and from the same NHL patients from whom we obtained the tissue biopsies (n=10) (**Figure 1A**, **Supplementary file 1: ST.1**). To profile 5mC/5hmC methylomes and matched gene expression activities, we extracted genomic DNA and RNA concurrently from the same samples, subjected the DNA to bisulfite (BS-seq) and oxidative bisulfite (oxBS-seq) whole genome sequencing (WGS) [55] and the RNA to RNA-seq (**Figure 1A**). In oxBS-seq, the DNA is first oxidized, which only converts 5hmC into 5-formylcytocine (5fC) followed by standard bisulfite treatment to convert both the 5fC and unmodified cytosine to T, leaving only 5mC protected. Therefore, sequencing the oxBS reactions yields the true 5mC sites. In a parallel bisulfite conversion reaction in the absence of the oxidation step, both 5mC or 5hmC are protected and called after sequencing. The 5hmC bases are then specifically identified by quantitative subtraction of the methylation levels determined between the oxBS-seq and standard BS-seq data (**Figure 1B**). Overall, high-quality and high coverage sequencing data were generated from each sample with high bisulfite conversion rates (**Supplementary file 1: ST.2**). On average, 58X and 47X BS-seq and oxBS-seq WGS data were produced, respectively with a mean bisulfite conversion rate of 98.6% (range, 97.04%-99.5%). Because cytosine methylation in mammalian cells occurs predominately at the CG dinucleotide context [56], we restricted our analyses to cytosines in the CpG context and methylated cytosines were defined as previously recommended by [44] (see Methods).

**Figure 1.**
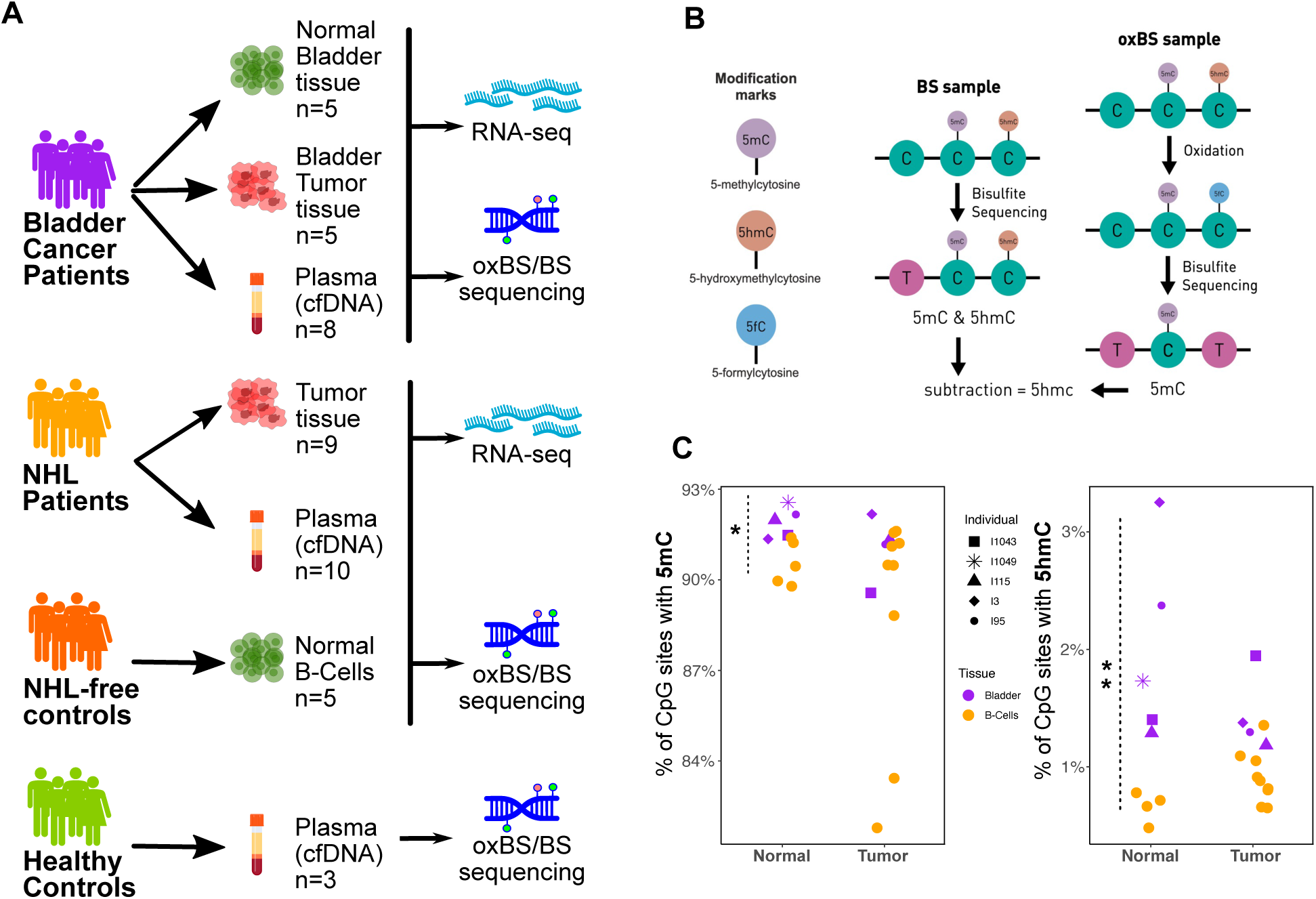
Global methylation analyses of tumor biopsies and paired healthy control from urothelial and lymphoid tissues. **A)** Schematics of the study design and patient cohorts. **B)** OxBS/BS sequencing strategy for the identification of 5mC and 5hmC at the single base resolution. In BS-seq, both 5hmC and 5mC are protected from bisulfite conversion while in oxBS-seq, 5hmC is first oxidized and converted to a 5-formylcytocine. From here, standard bisulfite conversation converts both the 5fC and unmodified cytosine to T and leaves 5mC untouched. To obtain 5hmC, the methylation levels from the oxBS-seq data are subtracted from the levels of the BS-seq data. **C)** Genome-wide levels of 5mC (left) and 5hmC (right) detected for all tissue samples represented as % of methylated CpGs. Significant differences dare labelled: * *p-value*<0.05; ** *p-value*<0.01. Note dots shapes represent different individuals only for bladder samples.

As expected, we observed high percentages of Cs in CpG context 5mC methylated across the genomes in all samples surveyed. On average, 91.9% Cs in CpG contexts (range, 91.4%-92.6%) and 90.6% (range, 89.8%-91.4%) were 5mC modified in Normal Bladder and Normal B-Cells, respectively, with a moderate but statistically significant higher 5mC in the former (2-group Mann-Whitney U Test, 5mC p-value = 0.01587). Meanwhile no statistically significant change of global 5mC level was detected when comparing to the global 5mC levels detected in cancerous tissues (Bladder Tumor vs. Bladder Normal: p-value=0.1905; NHL Tumor vs. Normal B-Cells: p-value=0.2366) (**Figure 1C**, **Supplementary file 1: ST.3**). In contrast to the 5mC’s abundance, 5hmC was only detected at 1-3% of total CpG, an order of magnitude lower than the levels (∼22%) previously surveyed in brain tissues and embryonic stem cells [57] (**Figure 1C**). Between Normal Bladder cells and Normal B-Cells, 5hmC levels varied substantially: bladder cells had a mean of 2.0% (range, 1.3%-3.3%) of Cs in CpG contexts with 5hmC while B-Cells had only 0.7% (range, 0.5%-0.8%) (2-group Mann-Whitney U Test, p-value= 0.007937) (**Figure 1C**, **Supplementary file 1: ST.3**). Similar to 5mC, global 5hmC levels were also found not significantly different between the malignant states and their matching normal (**Figure 1C**). Therefore, despite the reduction of 5hmC levels that has been reported in selective cancer types [12, 14, 15], we do not observe a global downregulation of 5hmC associated with oncogenic transformation.

### 5hmC is enriched at transcriptional enhancers and associated with tissue-specific gene expression

Across all tissue biopsies (n=23) surveyed by oxBS-seq and BS-seq, >90% of the CpG sites are methylated and the majority of 5mC sites exhibit a highly saturated methylation pattern with 57% of these loci having > 80% methylation frequency (i.e. the percentages of DNA molecules with methylated C per CpG locus). In contrast, 5hmC is highly heterogenous and often partial. At any given 5hmC site, only ∼30-40% of the DNA molecules surveyed are hydroxymethylated (**Figure 2A**). Furthermore, majority (∼ 97%) of CpG sites with 5mC occurs in a symmetric CpG dyad while 5hmC displays an asymmetrical strand bias. Such a hemi-modified 5hmC pattern can arise either through a partial 5mC demethylation or a *de novo* hydroxyl-methylation. We therefore investigated the relationship between 5mC and 5hmC and found a significant association between the presence of 5hmC and 5mC. Sites with 5hmC are more likely to have 5mC (Chi Square Test = 297637.6; p<1.0e-5) and, when 5mC and 5hmC are both detected on the same positions, their levels are negatively correlated (averaging between -0.53 to -0.84; **Supplementary file 2: Figure S1**). The median 5mC level is significantly lower (2-group Mann-Whitney U Test, 5mC p-value < 2.2e-16) for cytosines with 5hmC and vice versa (**Supplementary file 2: Figure S2A** and **S2B**). The observed correlations are consistent with the prior understanding that 5hmC, at least partially, is derived by 5mC oxidation [58, 59].

**Figure 2.**
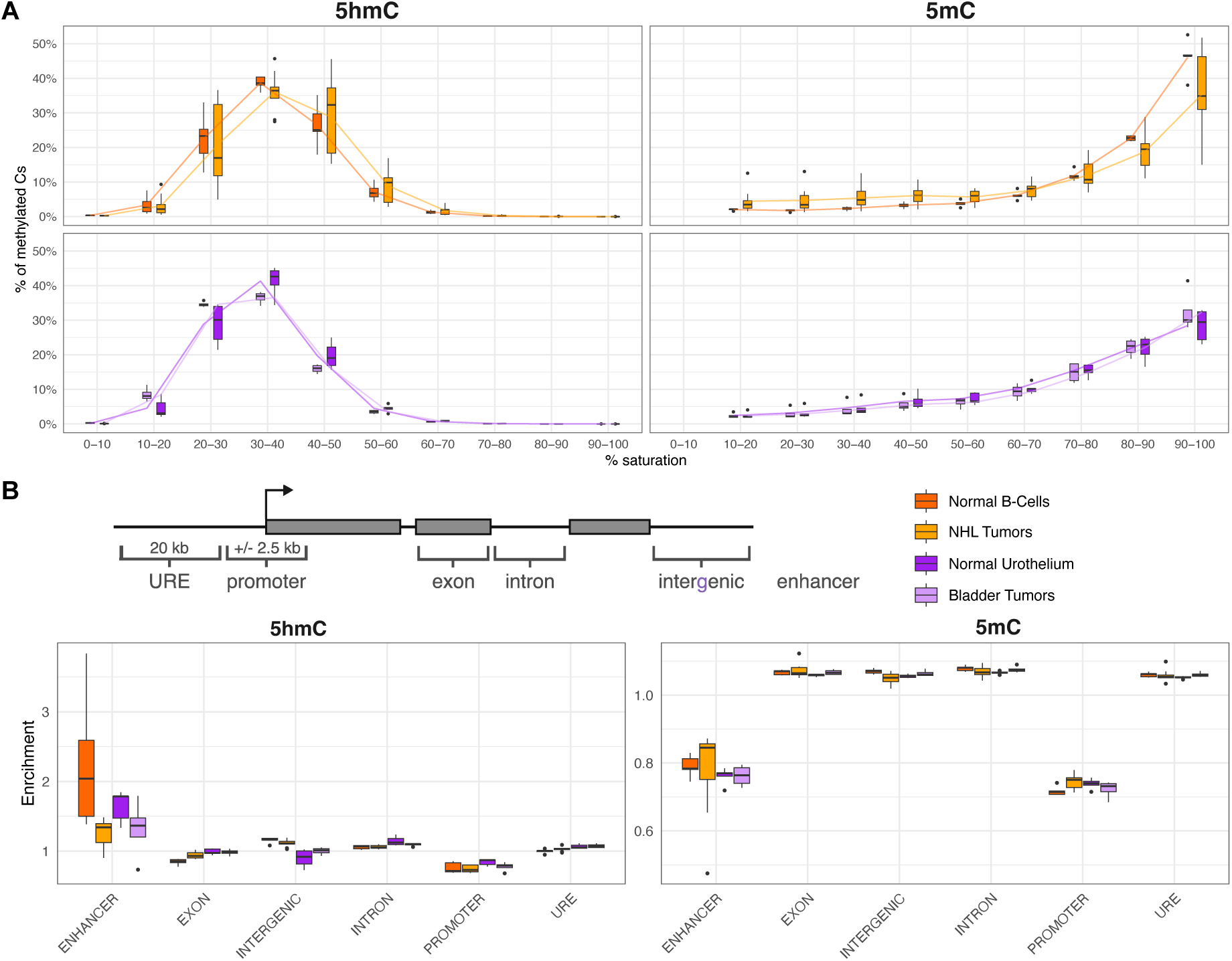
5mC and 5hmC methylation frequencies and genomic distributions. **A)** The percentages of methylated CpGs (Y-axis) detected at different saturation levels (X-axis). **B)** Distribution of 5hmC and 5mC across different genomic regions. The folds of enrichment at different regions are plotted for each sample type.

In addition to their negative correlations at sites where both 5mC and 5hmC are detected, 5mC and 5hmC exhibit distinct genomic distributions (**Figure 2B**). 5mC is uniformly distributed across most of the genomic regions including introns, exons, upstream regulatory elements (URE) (20Kb 5’ of the transcript start sites, TSSs) and intergenic regions while moderately depleted in promoters (±2.5Kb of the TSSs) and enhancer regions, consistent to its function as an epigenetic repressive mark. 5hmC, in contrast, is specifically enriched (mean=1.2-fold, range, 0.47-3.8) at enhancers (**Figure 2B**). Here, enhancers were defined as the 16,430 and 15,904 non-promoter histone H3 lysine 27 acetylation (H3K27ac) modified regions (FDR < 0.01) detected by ChIP-seq (see Methods) in the urothelial and germinal B-cells, respectively (**Supplementary file 1: ST.4**). Such 5hmC’s specific affinity to enhancers and regulatory regions presumably suggests a possible *de novo* hydroxymethylation in these regions. Interestingly, 5hmC levels in enhancers were lower in tumors compared with healthy tissues (2-group Mann-Whitney U Test, p-value= 0.03033) (**Figure 2B)**, making 5hmC of great interest when examining its regulatory function in oncogenic processes.

To further understand the functional significance of 5hmC, we sought to determine its tissue-specific patterns. 5hmC signal intensities are distinctive between Normal Bladder cells and Normal B-Cells; sample positioning along PCA’s PC1 (**Supplementary file 2: Figure S3**) suggest more similar methylation profiles among individuals from the same tissue than from different tissues. Such tissue-specific patterns were observed at coding genes and enhancer sites. We next performed differential methylation analyses (DMA) and identified 1,093 significantly differentially methylated genes (DMGs) (padj<0.05, FC>|2|) (**Figure 3A**, **Supplementary file 1: ST.5**). 717 and 376 genes exhibited 5hmC hypermethylation in bladder urothelium and B lymphocytes, respectively (**Figure 3B)**. Notably, when we further examined the nature of these 376 genes preferentially hypermethylated in germinal B-cells using EnrichR through Human Gene Atlas from BioGPS [53], these genes were significantly expressed in lymphocytes including CD19+, CD4+ and dendritic cells (3-5-fold, padj=2.7e-03 to 1.4e-05) (**Supplementary file 1: ST.6**), highlighting the tissue-specific nature of the 5hmC signatures.

**Figure 3.**
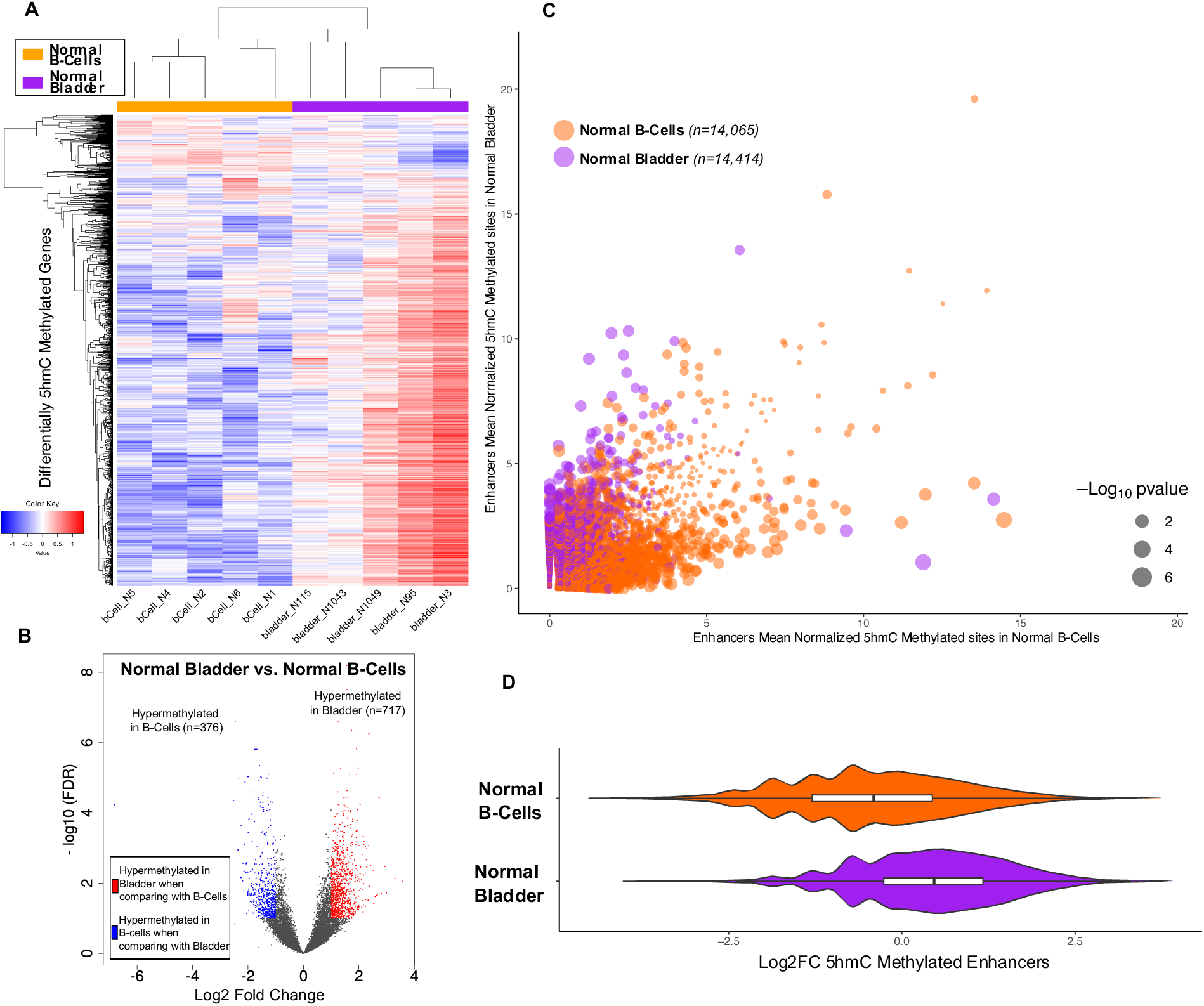
5hmC’s tissue specific modifications on genes and enhancers. **A)** The normalized methylation levels of differentially 5hmC methylated genes (DMGs) detected in Normal Bladder and Normal B-Cells displayed in a heatmap. **B)** Volcano plot showing differentially methylated genes between Normal Bladder tissue vs. Normal B-Cells. Blue dots indicate statistically significant hypermethylated genes in Normal B-Cells when comparing against Normal Bladder samples. Red dots indicate significant hypermethylated genes in Normal Bladder when comparing against Normal B-Cells. **C)** Scatter plot of the 5hmC methylation levels (normalized methylated sites) found in the enhancers from B-cells (orange) and bladder urothelium tissues (purple). Dot sizes represent the p-values from the differential methylation analysis. **D)** Log2 Fold Change for normalized 5hmC levels of the tissue-specific enhancers.

To evaluate the tissue specificity of 5hmC at enhancers, normalized 5hmC levels from the enhancer regions were compared between Normal Bladder cells and Normal B-Cells and found to be significantly elevated in their respective tissue types (Welch Two Sample t-test, t = 60.527, p-value < 2.2e-16, **Figure 3C** and **Figure 3D**). Through the DMA, we also identified 879 enhancers (DMEs) with differential levels of 5hmC, 531 hyper- and 348 hypo-methylated enhancers in Normal Bladder cells when compared their 5hmC levels to those found in Normal B-Cells (pval<0.05 & FC>|1.5|) (**Supplementary file 1: ST.7**).

To understand the potential impacts of these DMGs and DMEs on shaping the tissue-specific transcriptional programs, we examined gene expression by RNA-seq from the same tissue samples analyzed for methylation. High quality of RNA-seq data were generated and displayed good correlations among all 23 tissues (**Supplementary file 2: Figure S4**). Among the differentially expressed genes (n=10,606, padj<0.05 & FC>|2|) between Normal Bladder and Normal B-Cells samples (**Supplementary file 1: ST.8**), genes with elevated 5hmC levels are notoriously represented among genes upregulated in their corresponding tissues (**Figure 4A**, **Supplementary file 2: Figure S5**). Specifically, in Normal Bladder samples, genes with hypermethylated 5hmC have significantly higher RNA expression (mean log2 FC = 1.73, p-value = 4.5e-15, **Figure 4B**) while the hypomethylated genes (hypermethylated in B-Cells) show lower expression in Bladder (mean log2 FC = -0.49). Moreover, the fold-changes between 5hmC methylation and RNA expression display a moderate, but significant positive correlation (Pearson r=0.28, p-value < 2.2e-16), supporting the association between 5hmC modification and active transcription. The effects of 5hmC on transcriptional activation are also observed at genes associated with DMEs (**Figure 4D**). In total, there were 420 and 305 genes associated with hyper- and hypo-methylated enhancers in Normal Bladder cells, respectively when comparing to Normal B-Cells. These 420 genes proximal to enhancers with elevated 5hmC exhibit higher transcriptional activities (mean RNA-Seq log2 FC = 1.64) while the 305 genes proximal to the hypomethylated enhancers in bladder (hypermethylated in B-cells) are also downregulated in bladder (mean log2 FC = -1.29) (**Figure 4C**). Such differences in expression changes (1.64 vs -1.29) mediated by tissue specific 5hmC are highly significant (Welch Two Sample t-test =-13.1, p-value = 2.2e-16). Moreover, 5hmC enhancer hypermethylation and nearby-gene overexpression are highly indicative to their tissue origins. The 141 genes associated with hypermethylated enhancers and statistically significantly upregulated in normal bladder tissue are significantly enriched in bladder-specific pathways (*p*-value=2.8e-07, **Figure 4E**) while the 111 genes associated with hypermethylated enhancers, and statistically significantly upregulated genes in B-cells are significantly enriched in cytokine and immune-specific pathways (*p*-value=5.7e-05, **Figure 4F**).

**Figure 4.**
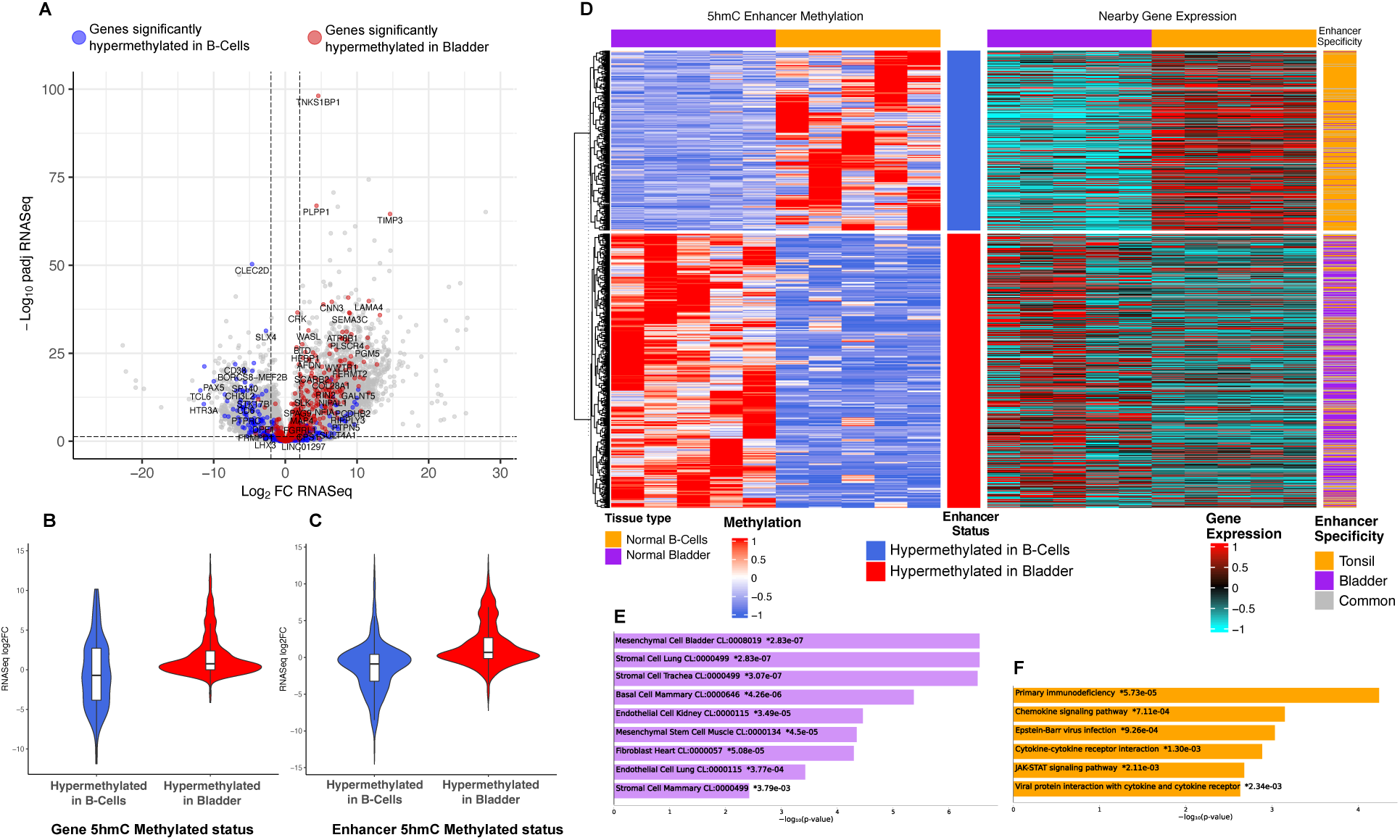
5hmC is associated with tissue-specific gene expression. **A)** Differential expressed genes in the Normal Bladder vs. Normal B-Cells comparison shown in a volcano plot. Genes with positive Log2 FC RNASeq (X-axis) are overexpressed in bladder and genes with negative values are overexpressed in B-cells. Genes with significantly differential 5hmC levels are colored in blue (hypermethylated in B-cells) and red (hypermethylated in bladder). **B/C**) Log2 Fold Changes (Y-axis) of expression from genes with differentially 5hmC methylated genes **(B)** and enhancers **(C)** showed in violin plots. X-axis shows methylated status for genes and enhancers. Positive values in the log2 Fold Changes of expression (RNASeq log2FC) indicate genes are overexpressed in bladder when comparing to B-cells, and vice versa. **D)** Heatmaps showing 5hmC normalized and standardized (z-scores) methylation levels from each significantly differentially methylated enhancers (left) and the normalized and standardized (z-scores) expression of the corresponding nearby genes (right) in Normal Bladder cells compared to Normal B-Cells. **E/F**) Enriched cell types and pathways from differentially expressed genes associated with 5hmC significantly hypermethylated enhancers in Normal Bladder (**E**) and Normal B-Cells **(F**).

### Differential 5hmC analysis reveals markers preferentially expressed in tumor biopsies

The comparative analyses of genome-wide 5hmC methylomes between distinct tissue types show that the 5hmC abundance in coding regions and enhancers reflects tissue-specific transcription. We reason that a systematic characterization of cancer-specific redistribution of 5hmC, with a focus on the genes and enhancers will shed light into new cancer markers. To uncover tumor-associated 5hmC changes, we applied the same strategies used in the normal tissue-based analyses and compared the 5hmC levels on gene coding and enhancer regions between cancer cells and their matched controls. 52 and 18 DMGs (padj<0.1 & FC>|1.5|, **Supplementary file 1: ST.9**) have differential 5hmC levels in Bladder Tumors and NHL Tumors samples, respectively. Consistent with the observed association between 5hmC signals and enhanced transcription, the 52 DMGs found in bladder cancers also exhibited significantly differential expression in Bladder Tumors when comparing to Normal Bladder (healthy cells) (p-value = 1.7e-0.2, **Figure 5A**). 5hmC hypermethylated genes in Bladder Tumors (n=9) are transcriptionally upregulated while the hypomethylated genes (n=43) are downregulated (mean RNA expression log2FC= 0.25 vs -0.54, respectively). Intriguingly, 14 out of the 18 DMGs found in NHL Tumors are located within ∼ 200 Kb region on chromosome 5 (chr5: 141,330,571-141,512,981) encoding the protocadherin gamma (*PCDHG)* gene clusters. The significance of this differential methylation region (DMR) in NHL Tumors is unclear and could be related to selection of normal controls (germinal center B-cells from reactive tonsillar tissues) used in the comparative analyses. Hence, they were excluded from subsequent investigations.

**Figure 5.**
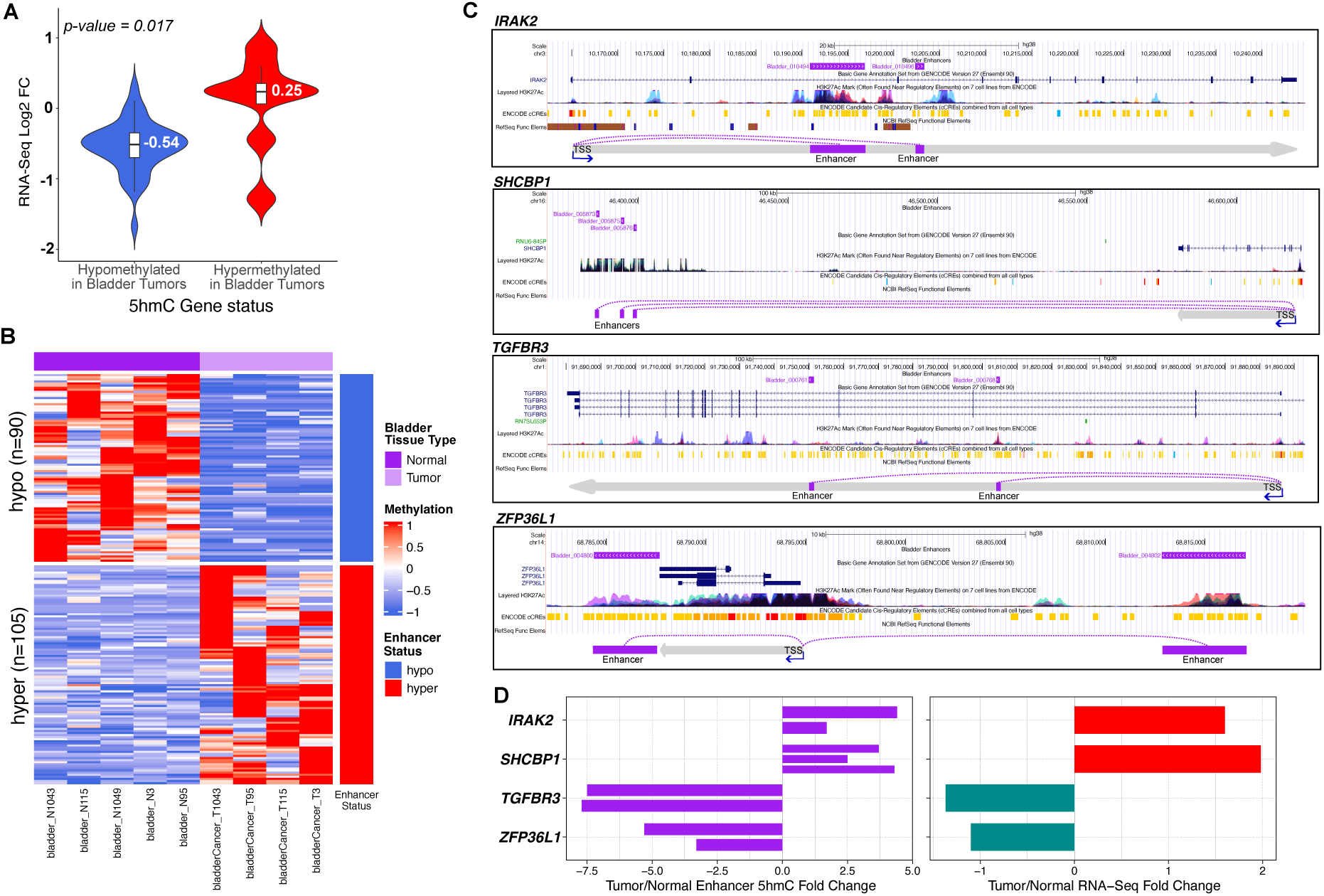
Differential 5hmC analysis reveals markers preferentially expressed in tumor biopsies. **A)** Violin plots showing RNA-Seq log2 FC values (Y-axis) from the 5hmC significantly differential methylated genes in Bladder Tumors when comparing to matched adjacent Normal Bladder tissues. **B)** A heatmap showing the normalized methylation levels of the differentially methylated enhancers detected in bladder tumors when comparing to normal bladder tissue. **C)** UCSC genome browser screenshots of regions harboring multiples DMEs and their affected genes detected in Bladder Tumor samples. The locations of the DMEs (purple) and transcription start sites (TSSs) of the genes (grey) are shown. Public annotated cis-regulatory element (CRE) and H3K27ac signals tracks from ENCODE are listed in reference. **D)** Fold Changes (X-axis) in 5hmC methylation values (Bladder Tumor vs. Normal Bladder) at DMEs (left) and the gene expression (right) of their corresponding affected genes (Y-axis) *IRAK2*, *SHCBP1*, *TGFBR3* and *XFP36L1* are shown.

When we examined the 16,430 urothelial enhancers in Bladder Tumor samples and compared their normalized 5hmC levels to Bladder Normal control samples, we observed 195 DMEs, including 105 hyper- and 90 hypo-methylated enhancers (*p*-value < 0.05 & FC > |1.5|, **Figure 5B**, **Supplementary file 1: ST.10**), and inferred 102 and 87 genes affected by these hyper- and hypo-methylated enhancers, respectively, based on their proximity to the nearest gene promoters. The expression of these genes as two distinct groups were not significantly up- or down-regulated in tumor tissues, presumably due to their multiplex enhancer regulation. We therefore focused on the genes affected by multiple DMEs. Genes targeted by >=2 hypermethylated enhancers, like *IRAK2* and *SHCBP1*, were expressed higher while genes targeted by multiple hypomethylated enhancers, like *TGFBR3* and *ZFP36L1*, displayed lower expression in bladder tumors (**Figure 5C, D**). Both *IRAK2* and *SHCBP1* are well known for their involvement in broad cancer types and their expression has been shown to drive cancer cell proliferation and *in vivo* tumor growth [60–66], while *TGFBR3* and *ZFP36L1*, both known as tumor suppressor genes, their inhibition can contribute to malignant transformation and induce tumor metastasis [67–70]. The concordant hyper- and hypo-5hmC intensities at their corresponding enhancers and their associated transcriptional activities suggest that the dynamics of 5hmC at the enhancers could be an indicator for transcriptional dysregulation in cancers. Specifically, the DMGs and genes affected by DMEs are highly enriched in the functional pathways relevant to tumorigenesis, like positive regulation of blood vessel endothelial cell migration (GO:0043536) (FDR<0.05), and many of them are known oncogenes and tumor suppressor genes like *EGFR, Suz12, AKT3* and *PPP2R5A*. 25 of the 52 5hmC DMGs and 34 from the 189 genes targeted by DMEs are known oncogenes defined in the Network of Cancer Genes resource (NCG) and The Catalogue Of Somatic Mutations In Cancer (COSMIC) [71, 72]. Taking *Suz12* as an example, *Suz12* exhibits the most hypermethylation (Fold Change=2.8) and its expression is strong in bladder tumor samples. As a subunit of polycomb complex, *Suz12*’s activation has been shown to predict tumor progression and aggressive characteristics in bladder cancer patients [73]. The most hypomethylated gene is *PPP2R5A*, whose expression is severely downregulated in bladder tumors. *PPP2R5A* encodes the Serine/threonine-protein phosphatase 2A and can act as a tumor suppressor to negatively control cell growth and division in cancer [74].

Taken together, our data suggest that the differential 5hmC levels can reflect tissue-specific transcription and aberrant gene expression associated with the oncogenic states.

### Characterization of 5hmC methylomes in cfDNA using a target-capture methylation assay

We reason that, if the methylation status in cfDNA represents the collection of DNA originating from all shredded cancer cells, then the 5hmC signatures found in tumor cells could be detected in the cfDNA from the patients’ plasma. The limited quantities of cfDNA that can be isolated from the human plasma present a challenge to achieve whole-genome, unbiased methylation profiling at sufficient coverages. Hence, we leveraged the findings from our comparative analyses of the tissue biopsies and adopted a scalable and effective SeqCap epi-target capture-based method [75] to evaluate if the 5hmC DMEs and DMGs uncovered in the tissue biopsies can be recapitulated in the plasma cfDNA of cancer patients. SeqCap targeted capture method profiles the 5hmC modifications observable in patients’ cfDNA through enrichment of fragments from pre-defined targeted genomic regions followed by BS/oxBS-seq to achieve higher sequencing coverages on regions of interests. To do so, we first designed a custom capture panel that includes genomic regions harnessing the DMGs and DMEs identified from the tumor-matched normal tissue comparisons. To enrich the contents and increase discovery power, we also include genome regions encoding known oncogenes and tumor suppressor genes from COSMIC’s cancer genome census and NCG6 Network of Cancer Genes [71, 72] where the primary sites or cancer types are bladder cancers or NHL. We incorporated genes that have known involvements in bladder cancers and NHL [76]. In total, this 30-megabases target capture panel includes high tiling densities of the probes from 348 genomic regions with total of 543,899 CpG sites (**Supplementary file 1: ST.11**).

Plasma cfDNA was collected from bladder cancer and NHL patients whose tumors were analyzed above and subjected to BS and oxBS conversion, target enrichment followed by amplification and sequencing. To confirm that the SeqCap epi-target capture approach generated reliable methylation status of the CpG loci within the regions of interests, we examined the correlations of the methylation levels between the whole-genome BS/oxBS-seq and targeted capture approaches in one of the samples and observed high correlation (Person correlation=0.93-0.94) (**Supplementary file 2: Figure S6A**). Moreover, high proportions of sequencing reads were found in the targeted regions (**Supplementary file 2: Figure S6B**, **Supplementary file 1: ST.3**), corroborating the high capture efficiency of the SeqCap approach. Using cfDNAs from bladder patients as a reference, good correlations (r=0.51, 0.53) of 5hmC levels were detected among different bladder cancer patients and between cfDNA and tumor tissues from the same patient with their distributions recapitulating the patterns found in the tissues (**Supplementary file 2: Figure S6C-D**). Overall, these quality metrices demonstrated the feasibility of the targeted capture approach to examine 5hmC on cfDNA and its resulted data quality.

### CfDNA recapitulates differential methylation genes detected in the tumor tissue biopsies

Using the SeqCap epi-target capture approach, we analyzed cfDNA samples from 8 bladder cancer patiens, 10 NHL patients and 3 healthy individuals (**Figure 1A**). Overall, 56% of total reads from the captured libraries were aligned to the targeted regions and on average 59% of the targeted cytosines in CpG contexts were subjected to 5hmC analysis (**Supplementary file 1: ST.3**). The overall 5hmC levels in the targeted regions were significantly lower in bladder cancer patients (2.7%) (Welch Two Sample t-test p-value < 0.0001) and NHL patients (3.4%) (Welch Two Sample t-test p-value = 0.0335) than in healthy individuals (5.8%), suggesting that the hypomethylated cfDNA could be an indicator of cancerous states. No significant change of total 5mC levels was detected in cancer patients’ cfDNA (bladder cancer patients = 85.3%; NHL patients = 84.9%; and healthy controls = 85.2%). To determine whether the differential 5hmC methylation observed in tissues can also be detected in cfDNA, the 5hmC levels from each of the candidate regions were normalized (see Methods) and subject to DMA. When cfDNA from patients of each cancer type were compared to the cfDNA from healthy controls, 100 and 33 differentially methylated regions (DMRs) (padj<0.1 & FC>|1.5|) were identified in plasma samples from patients with bladder cancers and NHL, respectively (**Supplementary file 1: ST.12**). Among the 100 DMRs in the cfDNA from bladder cancer patients, 30 were hypermethylated and 70 were hypomethylated (**Figure 6A**). Encouragingly, 69 of the 100 DMRs recapitulated the methylation patterns observed in bladder tissue biopsies (**Table 1**, **Supplementary file 1: ST.12**), including genes that were previously observed as DMGs in the bladder tumor biopsies like *AKT3*, *PPP2R5A*, and *FER* (**Figure 6B**). Similarly, among the 33 DMRs (6 hypermethylated and 27 hypomethylated) detected in the cfDNA from NHL patients (**Figure 6C**), 14 of them recapitulated the methylation patterns observed in neoplastic B-cells in NHL (NHL Tumors) (**Supplementary file 1: ST.12**), including genes like *ACTR3*, *FGFR3* and *PAPOLA* (**Figure 6D)**. Collectively, the DMRs with concordant tumor-associated 5hmC changes between tissues and cfDNA are summarized in **Table 1**.

**Figure 6.**
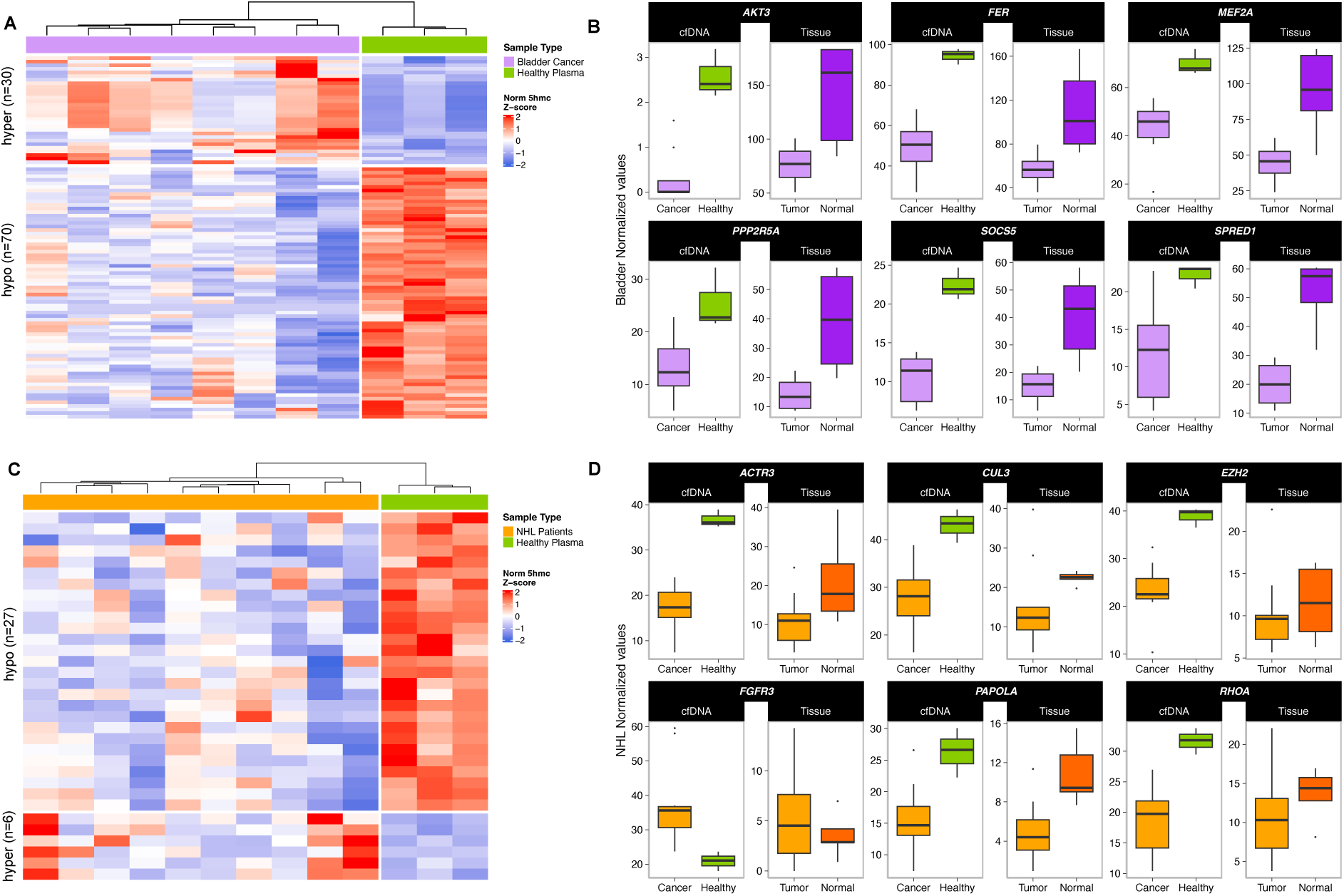
Methylation analysis of the DMGs detected in the tumor tissue biopsies in cfDNA. **A)** A heatmap showing the normalized 5hmC intensities of differentially methylated regions (DMRs) in the cfDNA from bladder cancer patients and healthy individuals. **B)** Examples of genes whose 5hmC levels displaying concordant downregulation in Bladder Tumors tissues and cfDNAs from bladder cancer patients. **C)** A heatmap showing the normalized 5hmC intensities of differentially methylated regions (DMRs) in the cfDNA from NHL patients and healthy individuals. **D)** Examples of genes whose 5hmC levels displaying concordant differential methylations in malignant B-cells (NHL Tumor tissues) and cfDNA from NHL patients.

**Table 1.**
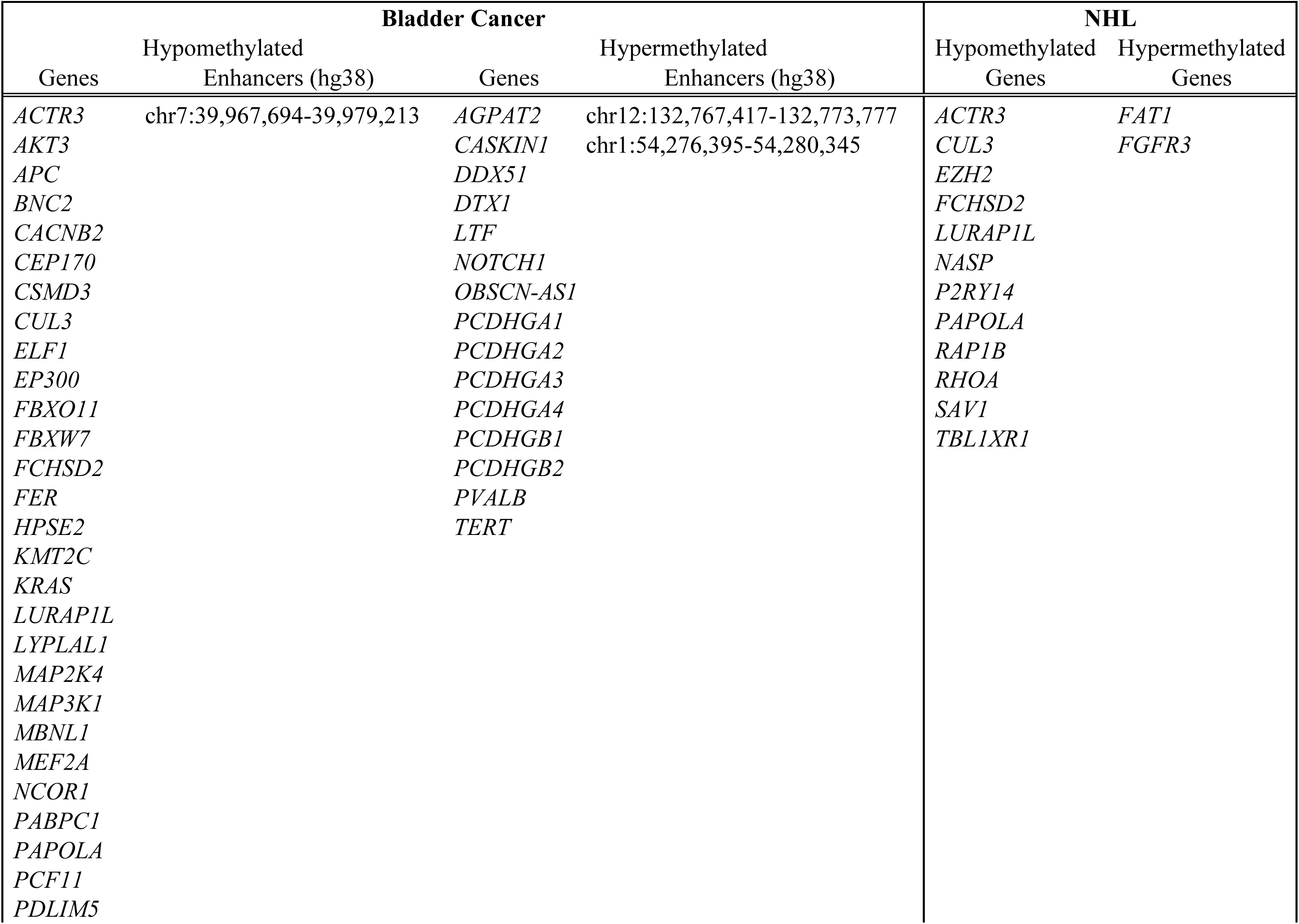
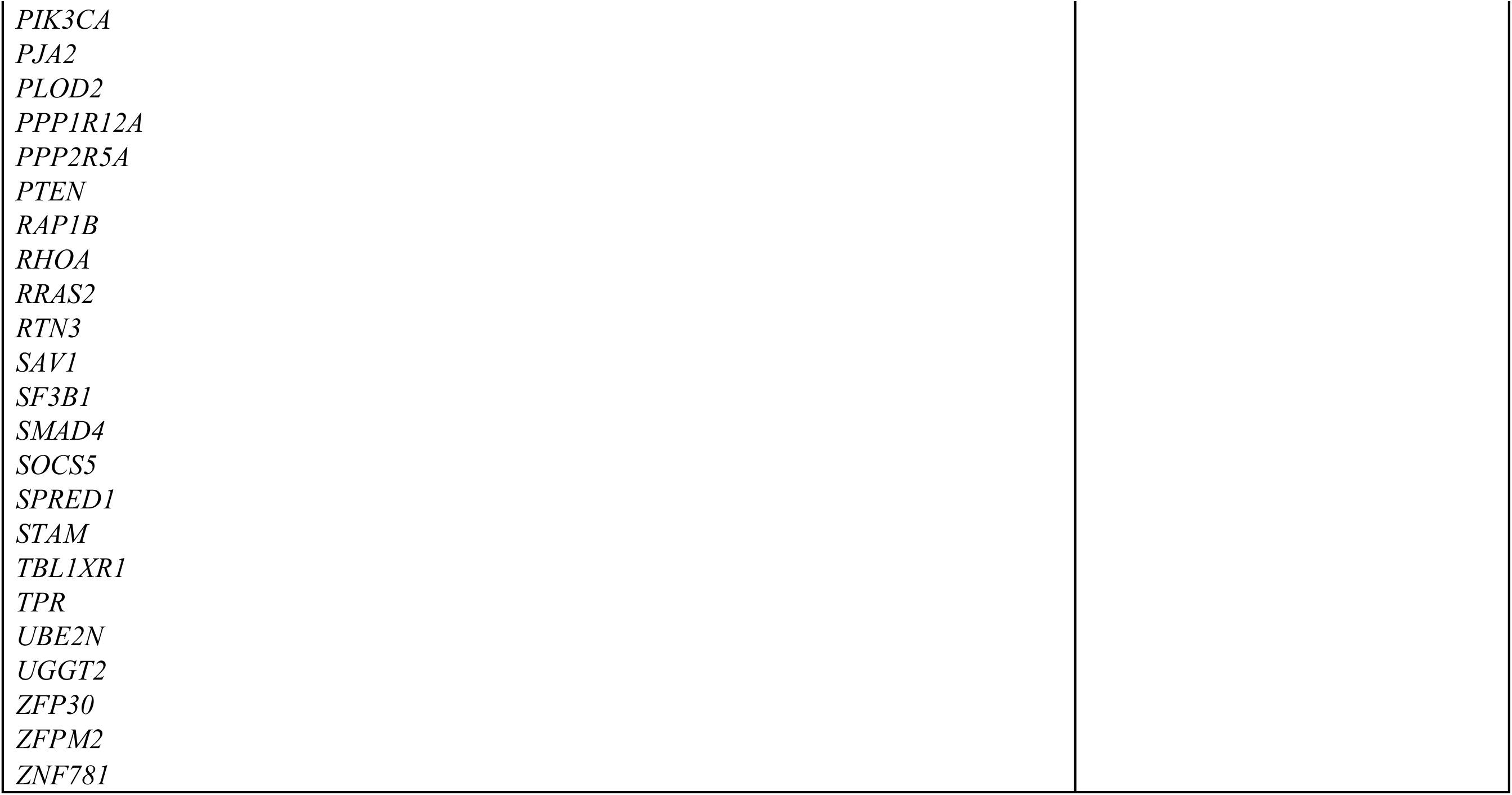
Summary list of DMRs showing differentially methylation when comparing cfDNA from plasma from cancer patients with healthy controls and showing a concordant pattern of methylation (hyper/hypo) in tissue samples.

| Bladder Cancer |  |  |  | NHL |  |
| --- | --- | --- | --- | --- | --- |
| Genes | Hypomethylated<br>Enhancers (hg38) | Genes | Hypermethylated<br>Enhancers (hg38) | Hypomethylated<br>Genes | Hypermethylated<br>Genes |
| *ACTR3* | chr7:39,967,694-39,979,213 | *AGPAT2* | chr12:132,767,417-132,773,777 | *ACTR3* | *FAT1* |
| *AKT3* |  | *CASKIN1* | chr1:54,276,395-54,280,345 | *CUL3* | *FGFR3* |
| *APC* |  | *DDX51* |  | *EZH2* |  |
| *BNC2* |  | *DTX1* |  | *FCHSD2* |  |
| *CACNB2* |  | *LTF* |  | *LURAP1L* |  |
| *CEP170* |  | *NOTCH1* |  | *NASP* |  |
| *CSMD3* |  | *OBSCN-AS1* |  | *P2RY14* |  |
| *CUL3* |  | *PCDHGA1* |  | *PAPOLA* |  |
| *ELF1* |  | *PCDHGA2* |  | *RAP1B* |  |
| *EP300* |  | *PCDHGA3* |  | *RHOA* |  |
| *FBXO11* |  | *PCDHGA4* |  | *SAV1* |  |
| *FBXW7* |  | *PCDHGB1* |  | *TBL1XR1* |  |
| *FCHSD2* |  | *PCDHGB2* |  |  |  |
| *FER* |  | *PVALB* |  |  |  |
| *HPSE2* |  | *TERT* |  |  |  |
| *KMT2C* |  |  |  |  |  |
| *KRAS* |  |  |  |  |  |
| *LURAP1L* |  |  |  |  |  |
| *LYPLAL1* |  |  |  |  |  |
| *MAP2K4* |  |  |  |  |  |
| *MAP3K1* |  |  |  |  |  |
| *MBNL1* |  |  |  |  |  |
| *MEF2A* |  |  |  |  |  |
| *NCOR1* |  |  |  |  |  |
| *PABPC1* |  |  |  |  |  |
| *PAPOLA* |  |  |  |  |  |
| *PCF11* |  |  |  |  |  |
| *PDLIM5* |  |  |  |  |  |

**Table 1.** Summary list of DMRs showing differential methylation when comparing cfDNA from plasma from cancer patients with healthy controls and showing a concordant pattern of methylation (hyper/hypo) in tissue samples.
|  |
| --- |
| *PIK3CA* |
| *PJA2* |
| *PLOD2* |
| *PPP1R12A* |
| *PPP2R5A* |
| *PTEN* |
| *RAP1B* |
| *RHOA* |
| *RRAS2* |
| *RTN3* |
| *SAV1* |
| *SF3B1* |
| *SMAD4* |
| *SOCS5* |
| *SPRED1* |
| *STAM* |
| *TBL1XR1* |
| *TPR* |
| *UBE2N* |
| *UGGT2* |
| *ZFP30* |
| *ZFPM2* |
| *ZNF781* |

In summary, these results suggest that tumor-associated 5hmC signatures can be detected in cfDNA. Together, these signatures can be used as potential features to distinguish cancer patients from healthy individuals. Our findings offer a molecular basis and exciting new opportunities to explore 5hmC as a biomarker in liquid biopsy-based applications.

## Discussion

In the present study, we analyzed DNA 5hmC and 5mC in paired tumor and matched normal tissues collected from bladder cancer patients as well malignant B-cells from germinal tonsils from NHL patients and normal B-cells from healthy individuals. Bladder cancer is the most common malignancy involving the urinary system [77, 78] and the NHL accounts for >10,000 deaths in the U.S. annually [79, 80]. Using a set of optimized technical and analytical methods, we determined genome-wide, single-nucleotide resolution 5hmC and its distributions in bladder tissues and germinal center naïve B-cells. Different from 5mC, 5hmC appears to be asymmetric, at low levels and enriched at tissue specific enhancers. Moreover, 5hmC signal displays broad variation at the same location across multiple samples, reflecting its dynamic nature. When comparing hydroxymethylation and gene expression, we demonstrate that the elevated 5hmC levels at genes and enhancers positively correlate with gene expression activity, further highlighting the role of hydroxymethylation in shaping the transcriptional programs. From paired bladder tumor and matched normal tissues, we comprehensively evaluated tumor methylomes and identified distinct 5hmC patterns. The discrete tumor-associated differential 5hmC is associated with expression changes and connected with pathways that are relevant in their respective tissues and cancer states, implicating that 5hmC is actively involved in epigenetic control, rather than an intermediate form of 5mC [81]. Finally, we showed that cfDNA is a faithful surrogate for tumor biopsy in identifying tumor specific methylation patterns. Surveying these targeted genes and regulatory elements opens exciting possibilities to the use of cfDNA methylome as a proxy of tumor epigenome for diagnosis, monitoring treatment and inferring tumor origins.

While this study provides insights into 5hmC’s functions in the transcriptional programs of malignant cells and its utility in a noninvasive genomic sequencing-based assay for cancer detection, future investigations are needed to address the current limitations and expand the value. Specifically, our finding is built on a limited number of samples and an expanded validation of the targets discovered here in larger patient cohorts will empower the specificity and sensitivity of the biomarkers in these cancer types. The minute quantity of cfDNA limits the ability to detect the broad types of genetic and epigenetic abnormalities associated with tumors. Considering the complexity of cancer-associated aberrations, technological advances in profiling multi-omic data at high sensitivity are needed to realize the potential of liquid biopsy for tumor detection. Several recent developments using more robust enzymatic treatment like Five-letter seq [82] and a duet evoC assay [83] which permits the unbiased and simultaneous detection of the genetic information as well as the 5mC and 5hmC modifications at base-resolution offer exciting promises in comprehensive analyses of cfDNA. Lastly, the nanopore single molecule sequencing offers capabilities in detecting multimodal analyses of cfDNA comprising of genetic alterations, DNA methylation and fragmentation patterns with benefit on shorter turnaround time, portability and lower costs [84–86]. Our study validates the concept and strategies for further assessing 5hmC values in clinical setting as well as providing initial insights into its pathogenic role. The study design and workflow devised here create an avenue that enables the broad applications to discover 5hmC biomarkers for tumors of different origins. Given its positive correlation with gene activities and enrichment in enhancers, 5hmC serves as a proxy for the oncogenic dysregulation that can be measured in cfDNA. In particular, the SeqCap Epi-target capture approach offers versatility and cost-effectiveness to validate targets from large cohorts and cross-reference the findings derived from other orthologous methods. The targeted capture and sequencing assays have been widely used as genomic testing to detect actionable mutations from multiple cancer types in clinical setting [87–89]. It is possible to incorporate 5hmC epialleles into targeted pan-cancer assays that offer broad utility in cancer detection and monitoring. Looking ahead, further validating these approaches in larger, more diverse populations of clinical subjects, is needed to prove its values in deploying epi-signatures as biomarkers in a liquid biopsy-based diagnostic test.

## Material and Methods

### Study design and sample collection

We employed a systematic approach to map genome-wide 5mC and 5hmC methylomes at single-nucleotide resolution using tumor biopsies and their matched normal tissues from a total of 23 samples across two distinct cancer cohorts: bladder cancer and non-Hodgkin lymphoma (NHL). For bladder, we obtained tumor and normal bladder tissues from 5 individuals with stage III (pT3) bladder cancer (**Figure 1A**). PT3 stage is defined as tumors exhibiting a pathological tumor-node-metastasis (pTNM), indicating that the invasive part of the tumor has grown through the muscle layers of the bladder into the surrounding fatty tissue layers. Normal tonsillar tissues (n=5) were obtained from routine tonsillectomy and germinal center B-cells were isolated by magnetic cell separation (CD38high, CD77+) as described [41]. Briefly, healthy tonsillar mononuclear cells were isolated by Ficoll-Hypaque density centrifugation and GC centroblasts (CD38^high^, CD77^+^) were stained with anti-CD77 (rat IgM isotype; Coulter), followed by incubation with mouse-anti-Rat-IgM (MARM), anti-IgG1 microbeads (Mitenyi Biotech) and isolated by magnetic cell separation. The cells were passed over an MS Column and the purity of the eluate was determined by staining against CD38, CD77 (revealed by MARM-FITC staining), and CD3. Patients with NHL undergoing excisional lymph node biopsies (n=9) were used to isolate neoplastic malignant B-cells by positive selection using CD19 Dynabeads (Life Technologies). Purity of the population was estimated by flow cytometry.

To analyze cfDNAs, paired blood samples were collected from the same NHL patients as the biopsies (n=10), from bladder cancer patients at the same pT3 stage (n=8), and from pooled plasma of healthy human donors (n=3) (**Figure 1A**) using anticoagulant K_2_EDTA from Innovative Research, Inc. (Novi, MI, USA). 10 mL of blood were collected in EDTA tubes and centrifuged at 800g for 15 min. Plasma supernatant was re-centrifuged at 3000 g for additional 15 min. The bladder cancer patient tissues and plasmas were obtained from the frozen biobank of the Research Tissue Repository Core Facility at University of Connecticut Health Center (Farmington, CT).

### DNA extraction

DNA was extracted using DNEasy kit (Qiagen) following manufacturer’s recommended protocol. cfDNA was isolated from 1 ml of plasma using the QIAamp® Circulating Nucleic Acid Kit (Qiagen). In Brief, plasma samples were thawed on ice and centrifuged at 4°C for 10 minutes at 16,000g and lysed for 30 minutes at 60°C using protease K and Qiagen lysis buffer. Supernatant was kept, and debris was discarded. Then the circulating nucleic acids were bound to the silica membrane of a QIAamp Mini column by applying vacuum pressure. After several washing steps to remove residual contaminates, the cfDNA was recovered from the membrane with an elution volume of 80 ul. The cfDNA was then quantified by Qubit dsDNA HS assay kit (Life Technologies) and the molecular size was determined with a 4200 TapeStation instrument and High Sensitivity D1000 or D5000 reagents (Agilent Technologies). The eluted cfDNA was stored at ≤ -20°C.

### Library preparation for BS- and oxBS-seq analysis

We adopted standard BS and oxBS sequencing approach. In brief, DNA libraries were constructed using the KAPA Hyper Prep Kit (Kapa Biosystems) following the manufacturer’s protocol. Each of the DNA libraries was resuspended with nuclease free water to a final volume of 49ul and subjected to oxBS and BS conversion treatment according to the manufacturer’s instructions described in the TrueMethyl Seq Kit v3.0 User Guide with the following modifications: unique indexes were incorporated for each library and PCR amplification cycles were reduced from 15 to 10. Treated DNA was cleaned up using bead-based purification and eluded in 25 ul of nuclease free water. Library quality was assessed using the 4200 TapeStation instrument and high sensitivity D1000 reagents (Agilent Technologies). DNA was quantified with Qubit dsDNA HS assay kit (Life Technologies). To monitor conversion efficiency, we added small amounts of synthetic methylation spike-in controls harboring DpnII restriction enzyme cutting sites with and without methylation. Bisulfite conversion rates were evaluated by the efficiency of DpnII digestion. Only libraries with high conversion rates were proceeded further for paired-end sequencing analyses. The oxBS-seq results in a direct readout of the 5mC profiles, while the 5hmC was determined by subtracting the oxBS-seq information from the BS-seq results (**Figure 1B**).

### 5mC and 5hmC calling

Adapters and low-quality bases were trimmed followed by alignment to the human reference genome GRCh38 primary assembly with bwa meth version 0.2.0 (Github: https://github.com/brentp/bwa-meth, [42]). PCR duplicates were marked using BISCUIT version 0.2.2.20170522 (https://github.com/zhou-lab/biscuit) [43]. At each cytosine location, we calculated the percentage of aligned bases that showed methylation (e.g. a cytosine aligned to the location) or no methylation (e.g. a thymine aligned to the location) using methyldackel extract version 0.3.0 (https://github.com/dpryan79/MethylDackel). To ensure high specificity in methylation calls, BS and oxBS C->T conversion efficiencies were determined from the cytosine site in the nonCpG context (calculated as total not methylated cytosines divided by the total number of cytosines sequenced). For all downstream analyses, only CpG sites were examined. CpG sites were defined using fastaRegexFinder.py version 0.1.1 against the reference assembly searching for “CG” with option “–no-reverse”. To avoid sex-based bias, we only analyzed methylation calls on the autosomes and all CpG sites that had >=10X coverage in both BS-seq and oxBS-seq were examined as recommended [44]. To assess the level of 5hmC at a CpG locus, we compared the numbers of retained cytosines in the BS-seq experiments to that in the oxBS-seq experiments. More specifically, we assumed that the number of retained cytosines follows a binomial distribution and performed a binomial proportions statistical test (using prop.test in R) in the oxBS and BS dataset, based on the proportion of reads containing a modified cytosine at that location to determine whether a cytosine is hydroxymethylated. 5hmC was calculated using custom R scripts as:

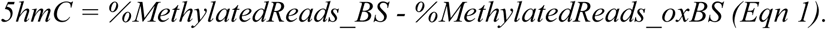

Any Cs with p>=0.05 were considered to have a 5hmC value of 0, and Cs with p<0.05 had a 5hmC value as calculated in equation 1 [45].

5mC was calculated as:

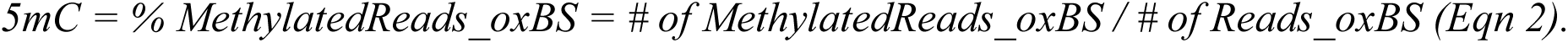

We consider 5mC to be present if >10% of the oxBS-seq reads aligning to a position within a CpG site show methylation. Details on the command used for the data processing and the required scripts are available on GitHub: https://github.com/gabyrech/tumorMethylome.

### Symmetry in Methylation Levels

All CpG dyads that had >=10x coverage in the BS dataset and oxBS dataset on both the C and G position were examined. CpG dyads with 5hmC > 0 on both the C and G are considered as “symmetric” for 5hmC. Similarly, CpG dyads with 5mC > 10% on both the C and G are considered as “symmetric” for 5mC. A CpG dyad is defined as “hemi” methylated if only the C or G position, but not both, is methylated. We then calculated the Spearman and Pearson correlations between the level of methylation on the C and G across all symmetric dyads.

### Methylation patterns in genomic regions

We defined genomic regions using Gencode’s annotation of the human reference genome (GRCh38 primary assembly, Gencode version 27), from which we retained only the following biotypes: protein_coding, lincRNA, antisense_RNA, miRNA, sense_intronic, processed_transcript, sense_overlapping, IG_V_gene, TR_V_gene, TR_J_gene, IG_D_gene, IG_J_gene, IG_C_gene, TR_C_gene, TR_D_gene, totalizing 35,412 genes. We defined promoter regions as +/-2.5kb of the transcription start sites (TSSs) and upstream regulatory elements (URE) as 20kb upstream of the TSSs. Exons were defined by the Gencode v27 annotation, and introns were defined as the space between the exons. The remaining positions were defined to be intergenic. The cytosines in CpGs were found across the genome with the following distribution: 18% in promoters, 2% in exons, 25% in introns, 26% in UREs, and 29% in the intergenic regions.

Enhancer annotations were defined for each tissue type [46–48]. For bladder tissue, a H3K27ac ChIP-Seq data from a human bladder tissue generated through the Encode resource [46] ENCSR054BKO (GSM1013133) was downloaded. For germinal B-cells, H3K27ac ChIP-Seq data [48] from GEO (accession: GSE89688) was downloaded. We performed MACS peak calling on these datasets using the methodology described [49] and retained only peaks with FDR < 0.01 on autosomes. 5hmC-modified enhancers were defined from the merged H3K27ac peaks (< 1Kb) outside of the promoters that displayed 5hmc modification from minimally one sample. To ensure sufficient signals for DMA, the H3K27ac peaks spanning less than 1 Kb were extended from the summits to up to 1Kb in length. The enhancer-associated target genes were defined by genes whose transcription start site (TSS) (according to Gencode v27 annotation) has the shortest distances to the corresponding enhancers using *bedtools closest*.

To determine the distributions of 5hmC and 5mC across genomic regions of different features, we compared the proportion of Cs in CpG sites with 5hmC (or 5mC) for a given genomic region type (e.g. intron, exon, etc.) to the average total proportion of Cs in CpG sites that have 5hmC (or 5mC). In the case of enhancers, we compare the proportion of Cs in CpG sites with methylation in enhancers to a bootstrapped background genome distribution matching the ratio at which the enhancers fall into genomic regions. The background distribution for each sample was created by sampling 5hmC (or 5mC) levels from 10,000 covered Cs spread across the genome following the same ratios as enhancers fall into genomic regions 500 times. From the distribution of 500 samples, the median # of Cs with 5(h)mc were used as the background to calculate enrichment.

### Differential methylation analysis (DMA)

To determine differentially methylated genes or enhancers, we first created a matrix with the total number of methylated Cs per region based on the 5mC and 5hmC calling detailed before. For gene-level analysis, we used Gencode v27, extending the coordinates to include 1 kb upstream of the TSSs. After the initial exploratory data analysis, we identified one of the Bladder Tumor samples (bladderTissue_T1049) as an extreme outlier when compared to all other tissue samples. Consequently, this sample was excluded from the DMA and all subsequent analyses. DMA was performed using DESeq2 [50], which employs a normalization model that internally corrects for sequencing depth / library size, making the samples comparable to each other. After normalization, DMAs were performed between different groups of samples. At the tissue level, in order to determine tissue-specific DMRs we compared Normal Bladder samples (n=5) vs Normal B-Cell samples (n=5). For identifying cancer-specific DMRs, we compared methylation levels between tumor samples and normal samples: Bladder Tumor (n=4) vs. Normal Bladder (n=5) and malignant B-Cells from NHL patients (NHL Tumors, n=9) vs. Normal B-Cells (n=5). Finally, at the cfDNA level, we compared methylation levels in cfDNAs from patients with bladder cancer (n=8) vs. healthy controls (n=3) and cfDNAs from patients with NHL (n=10) vs. healthy controls (n=3).

### Differential gene expression analysis

RNA was extracted using RNeasy kits (Qiagen) following manufacturer’s recommended protocol. RNA-Seq library prep was performed following the Poly(A) RNA-Seq – Kapa Stranded mRNA-seq kit protocol. Reads from RNA-seq data were trimmed to remove adapters and low-quality sequences using Trim Galore! (https://github.com/FelixKrueger/TrimGalore) with the following parameters: *--paired --stringency 3 --length 20 -q 30*. Trimmed reads were then aligned to Homo sapiens GRCh38 primary assembly with hisat2 (v.2.1.0) [51]. We used HTSeq [52] to quantify transcript abundance with parameters for reverse strandedness (-s=reverse), genes considered as the union of all its exons (--mode=union) and assigning reads that are assigned to more than one feature to all aligned featured (–nonunique=all). We used gene models from GRCh38 primary assembly, Gencode v27, but after quantification we kept only biotypes described above (35,412 genes). We used the matrix of gene counts to perform differential gene expression analysis with the R package DESeq2 [50]. Genes were considered differentially expressed between two groups when the adjusted p-value (padj) < 0.05 and fold change (FC) > |2|. Gene enrichment analysis for differentially expressed genes associated with 5hmC significantly differentially methylated enhancers were performed using EnrichR [53] based on Tabula Muris single cell transcriptome data (https://tabula-muris.ds.czbiohub.org/) for the 141 genes associated with hypermethylated enhancers in Bladder and on KEGG 2021 Human for the 111 genes associated with hypermethylated B-Cell enhancers.

### CfDNA methylation analysis

#### SeqCap epi-target capture panel

To enable higher coverages of genomic regions of interest from limited quantities of cfDNA, we created a custom 30MB capture panel covering genes and enhancers with evidence of being involved in the bladder cancer or lymphoma. In total, 348 features were selected which included known tumor suppressor genes and oncogenes from COSMIC’s cancer genome census and NCG6 Network of Cancer Genes where the primary site or cancer type is either bladder cancer or lymphoma. We also performed a thorough literature review and incorporated genes that have known involvement in bladder cancer or lymphoma.

#### Capture library preparation and 5hmC calls

High quality cfDNA was extracted, end-repaired and A-tailing. NimbleGen SeqCap adaptors (Roche) were ligated to the end-repaired DNA using ligation components in the KAPA HTP library preparation kits and then processed in parallel through both BS-seq and oxBS-seq library protocols. All libraries were hybridized to the custom SeqCap Epi probe panels using the standard SeqCap Epi workflow (Roche). Post-capture libraries were quantified by qPCR using KAPA library quantification kits, pooled and sequenced with non-bisulfite converted DNA as spike-in. Numbers of methylated cytosines per feature from each sample was determined in a similar fashion as previously used in tissue samples except 7X was used as the minimal required coverage in data from both BS-seq and oxBS-seq for a cytosine to be considered in the analysis.

### Ethics approval and consent to participate

This study was conducted in accordance with the principles of the Helsinki Declaration and were approved by the appropriate Institutional Review Boards: Oregon Health Science University (OHSU) IRB#4422 and IRB#4918, The Jackson Laboratory IRB Number 2018-025. All patients provided written informed consent.

### Availability of data and materials

All data described in this study can be found at the NCBI Short Read Archive, BioProject: PRJNA1184365. Computational methods and tools used in this study are listed in the Methods and can be downloaded from https://github.com/gabyrech/tumorMethylome. Further information and requests for resources and reagents should be directed to and will be fulfilled by lead contact Chia-Lin Wei.

## Acknowledgments

We gratefully acknowledge the contribution of the Genome Technologies Service at The Jackson Laboratory for expert assistance with the work described herein.

## SUPPLEMENTARY FILES

**Supplementary file 1. Supplementary Tables: Contains additional tables supporting the study analyses and results.**

ST.1. Summary of the patient information and tumor characteristics used in this study.

ST.2. Sequencing metrics of tissue samples subjected to bisulfite (BS-seq) and oxidative bisulfite sequencing (oxBS-seq).

ST.3. Summary metrics for 5mC and 5hmC calls from tissues and cfDNAs.

ST.4. Enhancers selected for 5hmC analysis in this study.

ST.5. 5hmC differentially methylated genes (DMGs) found in normal tissues (Normal Bladder cells vs. Normal B-Cells). Negative Fold Changes indicate genes hypermethylated in normal B-Cells when comparing against Normal Bladder cells. Positive Fold Changes indicate genes hypermethylated in Normal Bladder when comparing against normal B-Cells.

ST.6. Gene list enrichment analysis of the 376 5hmC hypermethylated genes in Normal B-cells when comparing with Normal Bladder tissues. Enrichment analyses was performed using EnrichR over the Human Gene Atlas available through BioGPS.

ST.7. Differentially 5hmC methylated enhancers detected in Normal Bladder and Normal B-Cells). Positive Fold Changes indicate enhancers hypermethylated in normal bladder when comparing against normal B-cells. Negative Fold Changes indicate enhancers hypomethylated in normal bladder when comparing against normal B-Cells.

ST.8. Differentially expression analysis for tissue samples: Normal tissues (Bladder vs. B-Cells); Tumor/Normal (NHL Tumors vs. Normal B-Cells, and Bladder Tumor vs. Normal Bladder).

ST.9. Differentially 5hmC methylated genes identified through comparisons of Bladder Tumor vs. Normal Bladder and NHL Tumors vs. Normal B-Cells. Note Fold Change (FC) always refers to tumor samples relative to their respective controls (normal samples).

ST.10. 5hmC differentially methylated enhancers detected by comparison between Bladder Tumor vs. Normal Bladder at the enhancer level. Note positive Fold Change indicates hypermethylation in tumor samples and negative indicates hypomethylation relative to normal bladder samples controls.

ST.11. Genes and enhancers included in the content of the targeted capture panel.

ST.12. Summary of the 5hmC differentially methylated regions (DMRs) identified in cfDNA by comparing cancer patients with healthy controls. Region Type: Whether the analyzed region is a Gene or an Enhancer. Gene Name: Name of the gene or closest gene of the enhancer. DMR annotation: either Hypomethylated or Hypermethylated in the cfDNA of cancer patients when comparing to healthy individuals. (*) means the difference is statistically significant, n.s.: not significant. Tissue 5hmC: Whether the methylation status between the tissues and plasma cfDNA are concordant.

**Supplementary file 2. Supplementary Figures: Contains additional figures supporting the study analyses and results.**

**Supplementary Figure S1.** Correlations between 5hmC and 5mC from all 23 tissue samples analyzed. Densities of all Cs in CpGs (>=10x coverage in BS-seq and oxBS-seq) with different methylation frequences were plotted. **Lower right**: Spearman correlations between 5mC and 5hmC for all tissue samples, split by sample type Normal Bladder, Bladder Tumors, Normal B-Cells and NHL Tumor samples.

**Supplementary Figure S2.** The levels of 5mC (**A**) and 5hmC (**B**) at CpG sites with both methylation markers and with only one respective methylation marker from different tissue types are shown.

**Supplementary Figure S3.** Principal Component Analysis (PCA) of normalized 5hmC and 5mC methylated sites at enhancers **(A)** and gene regions **(B)** for Normal Bladder and Normal B-Cells tissue samples.

**Supplementary Figure S4**. Pairwise correlations of normalized gene counts between all 23 tissues from RNA-Seq data.

**Supplementary Figure S5**. Volcano plot showing results from the differential expression analysis between Normal Bladder and Normal B-cells. Positive Log_2_ Fold Change in RNA-Seq (X-axis) indicate genes are overexpressed in Bladder, while negative values indicate genes are overexpressed in B-Cells. Genes are colored based on their differential 5hmC methylation Log2 Fold Change (Log_2_ FC 5hmC): positive (red) and negative (blue) values represent the hyper- and hypomethylation genes in Bladder when comparing to B-Cells, respectively.

**Supplementary Figure S6**. Quality assessments of the cfDNA 5hmC data generated by SeqCap epi-target capture analysis. **A)** Correlations of the methylation calls and levels (oxBS-Seq at the left and BS-seq at the right) between the whole-genome sequencing and targeted capture approaches using sample N3 (Bladder Normal). **B)** Visualization of the coverages and sequence reads aligned to the genomic region (chr5:108,095,545-109,848,119) harboring target regions (genes *FER* and *PJA2*) marked in green by integrated genome browser (IGV), BS-seq (top) and oxBS-Seq (bottom). **C)** Correlation of 5hmC levels between cfDNAs from different bladder cancer patients. **D)** Correlation of 5hmC between cfDNA and tumor tissue from the same individual and their 5hmC frequency distributions.

