## Supplementary file 2 for "Global DNA methylomes reveal oncogenic-associated 5-hydroxylmethylated cytosine (5hmC) signatures in the cell-free DNA of cancer patients"

### Supplementary Figure S1.

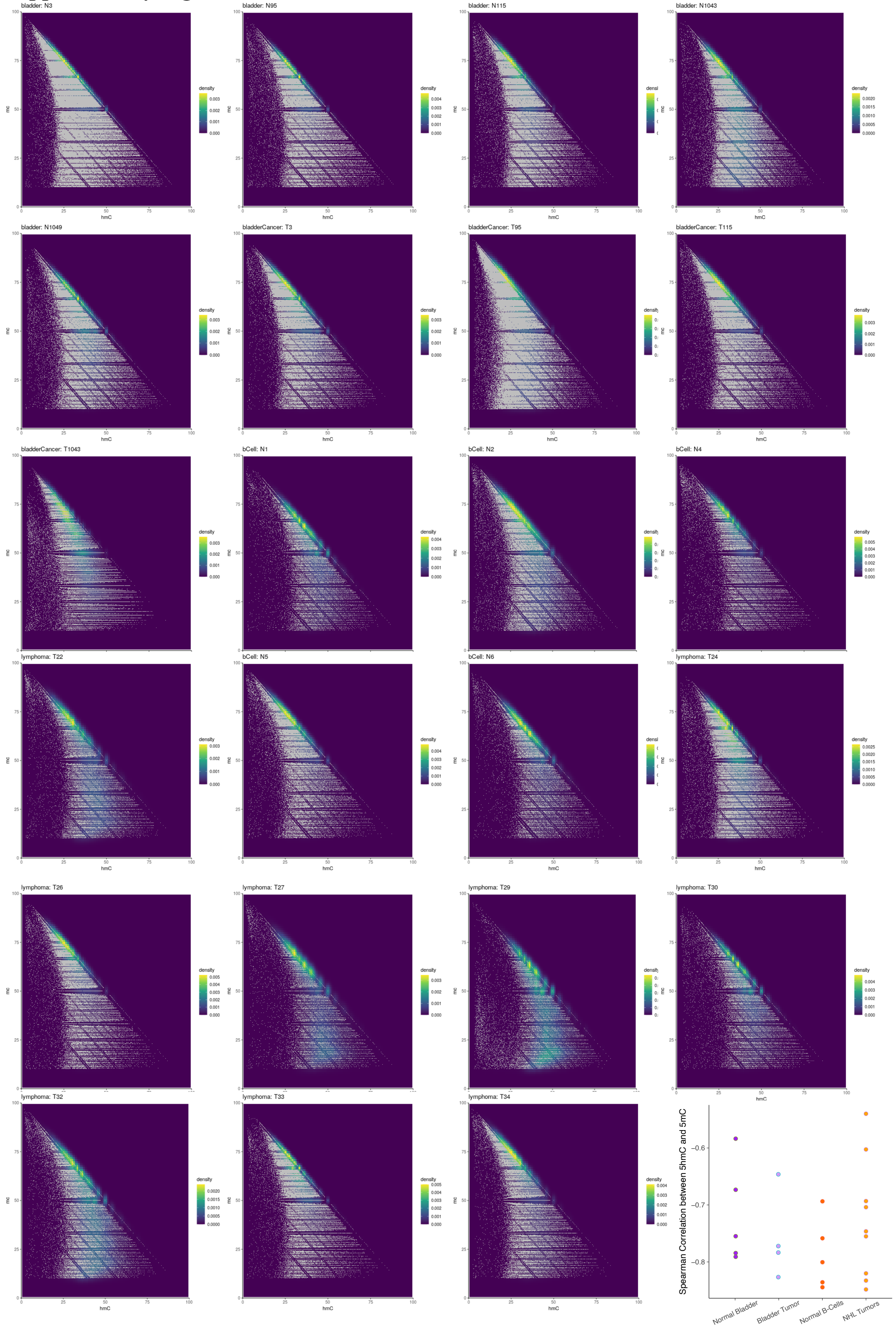

**Supplementary Figure S2.**

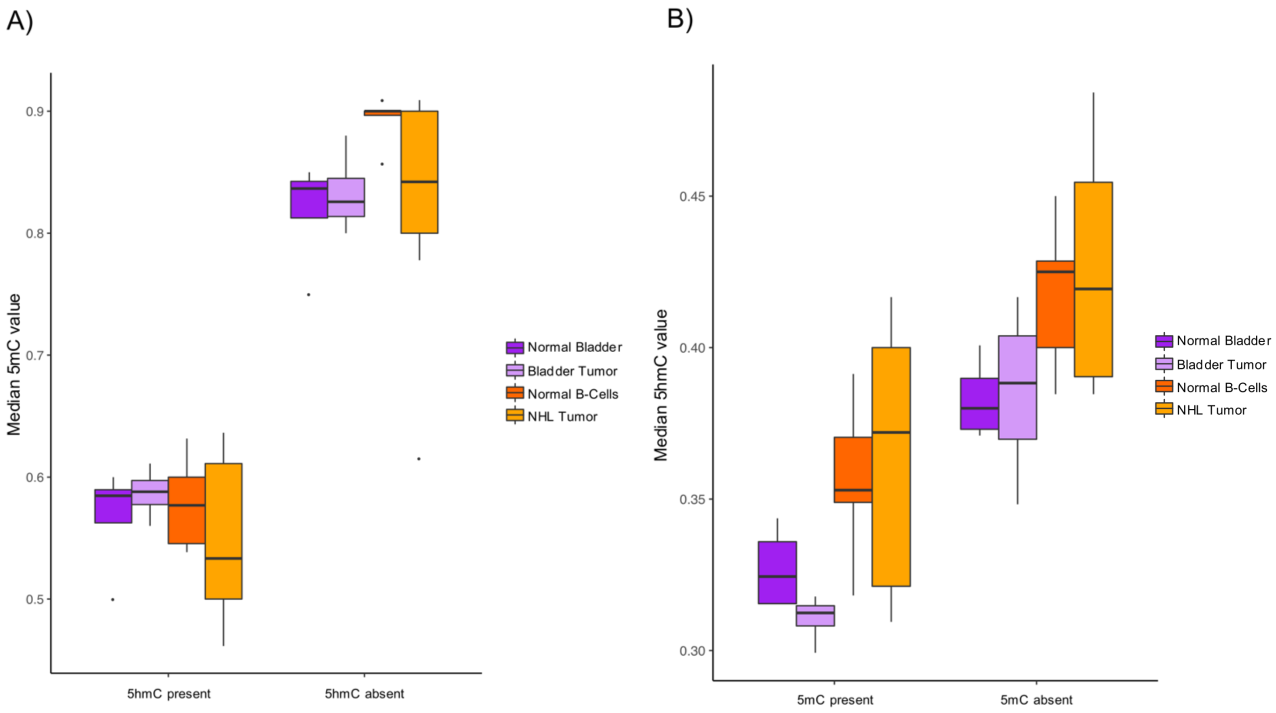

**Supplementary Figure S3.**

A)

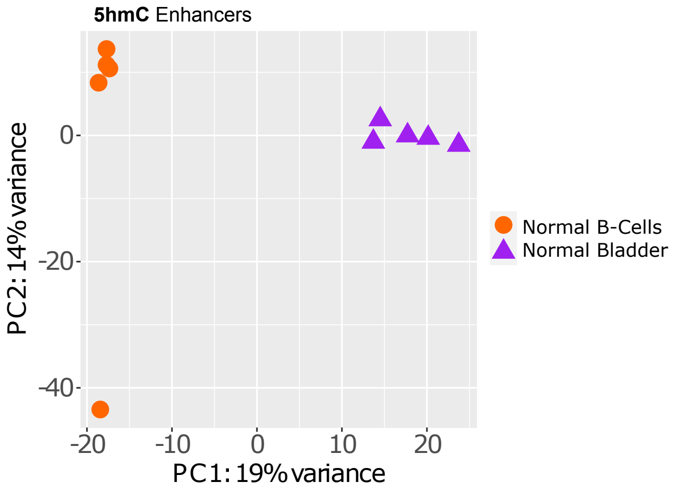


B)

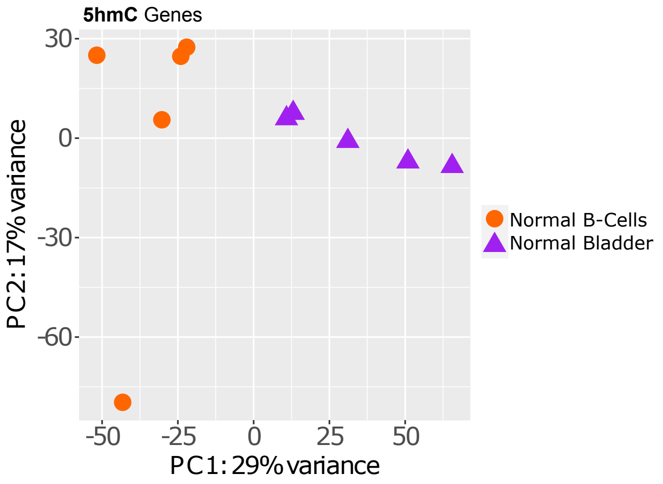

**Supplementary Figure S4**.

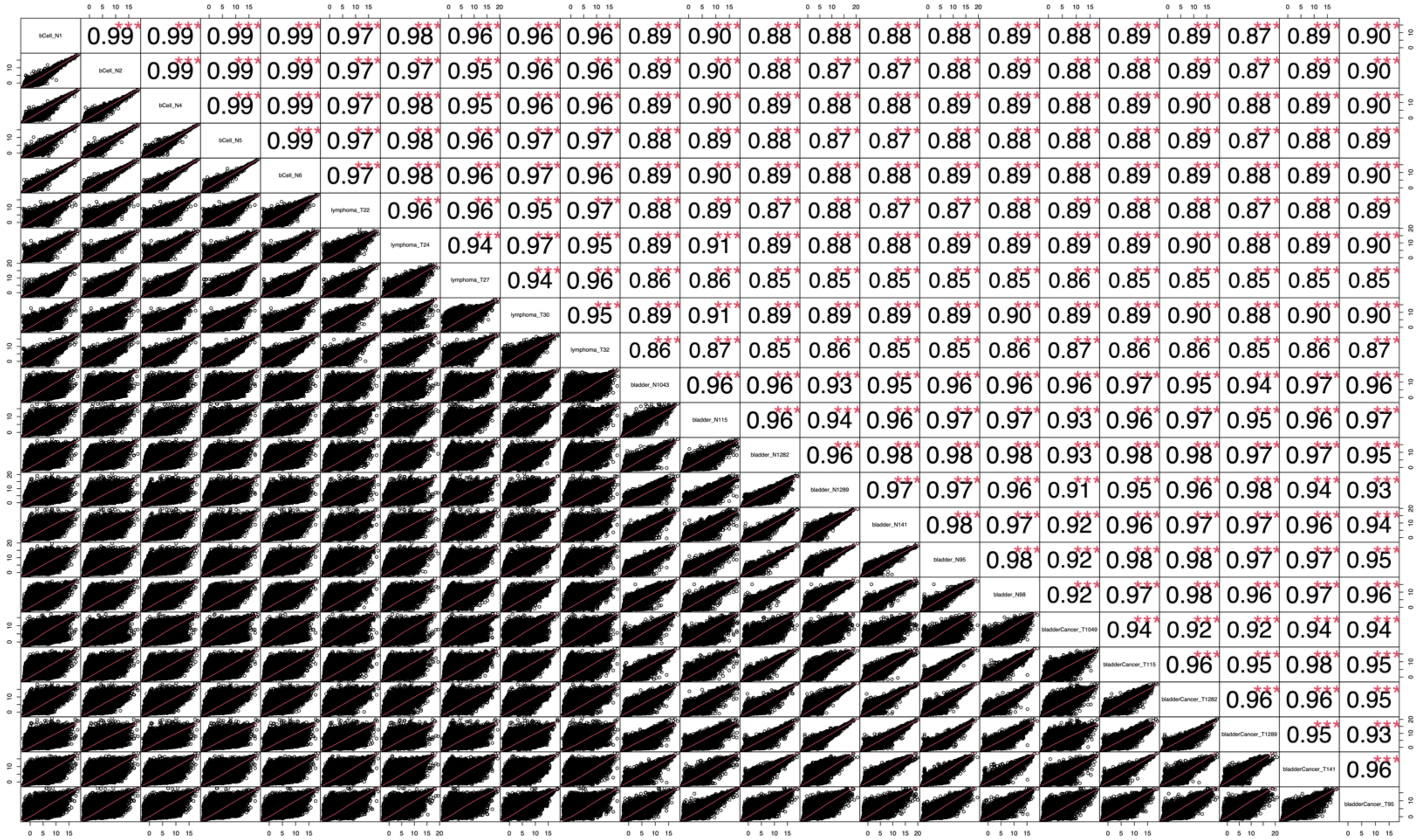

**Supplementary Figure S5.**

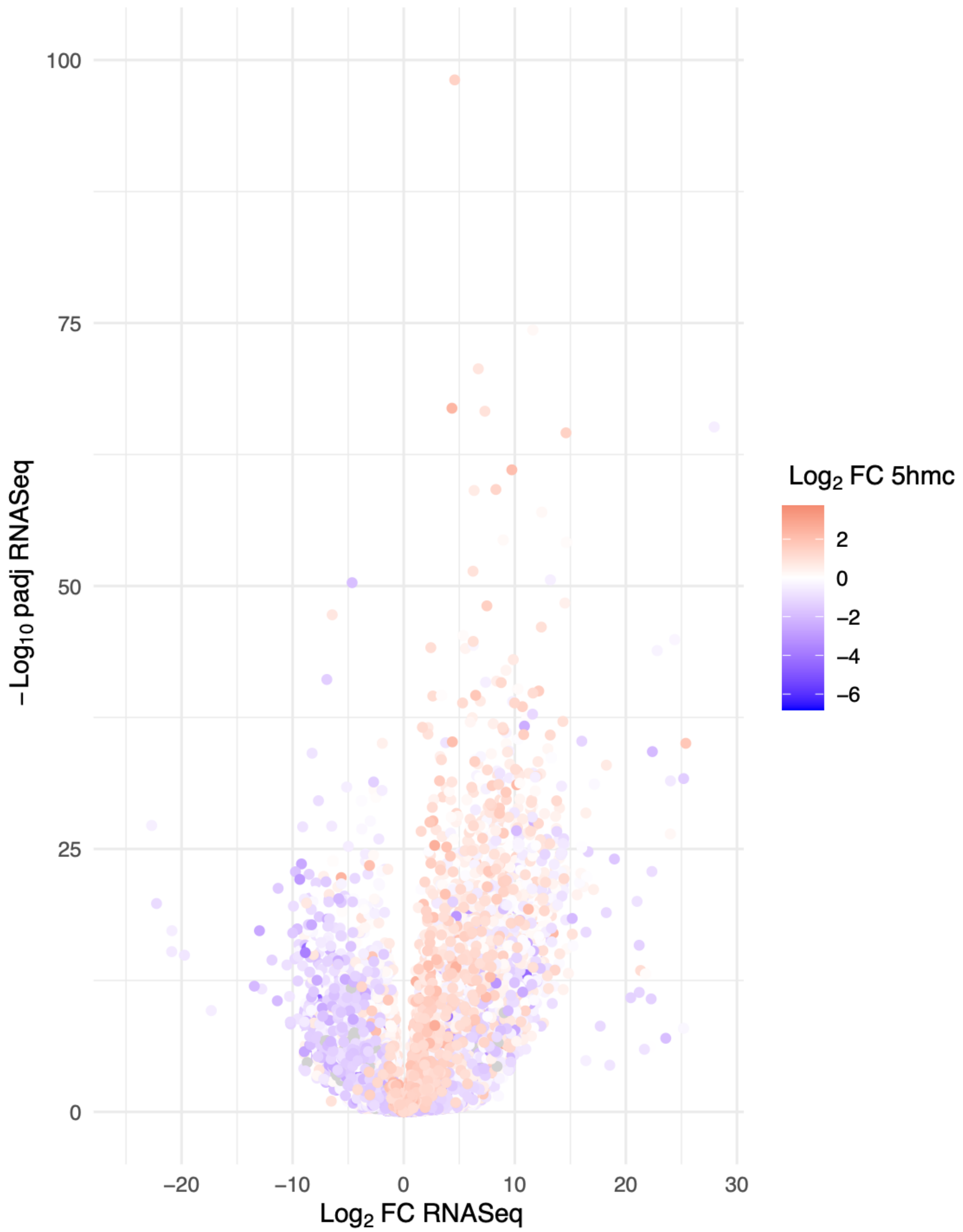

**Supplementary Figure S6.**

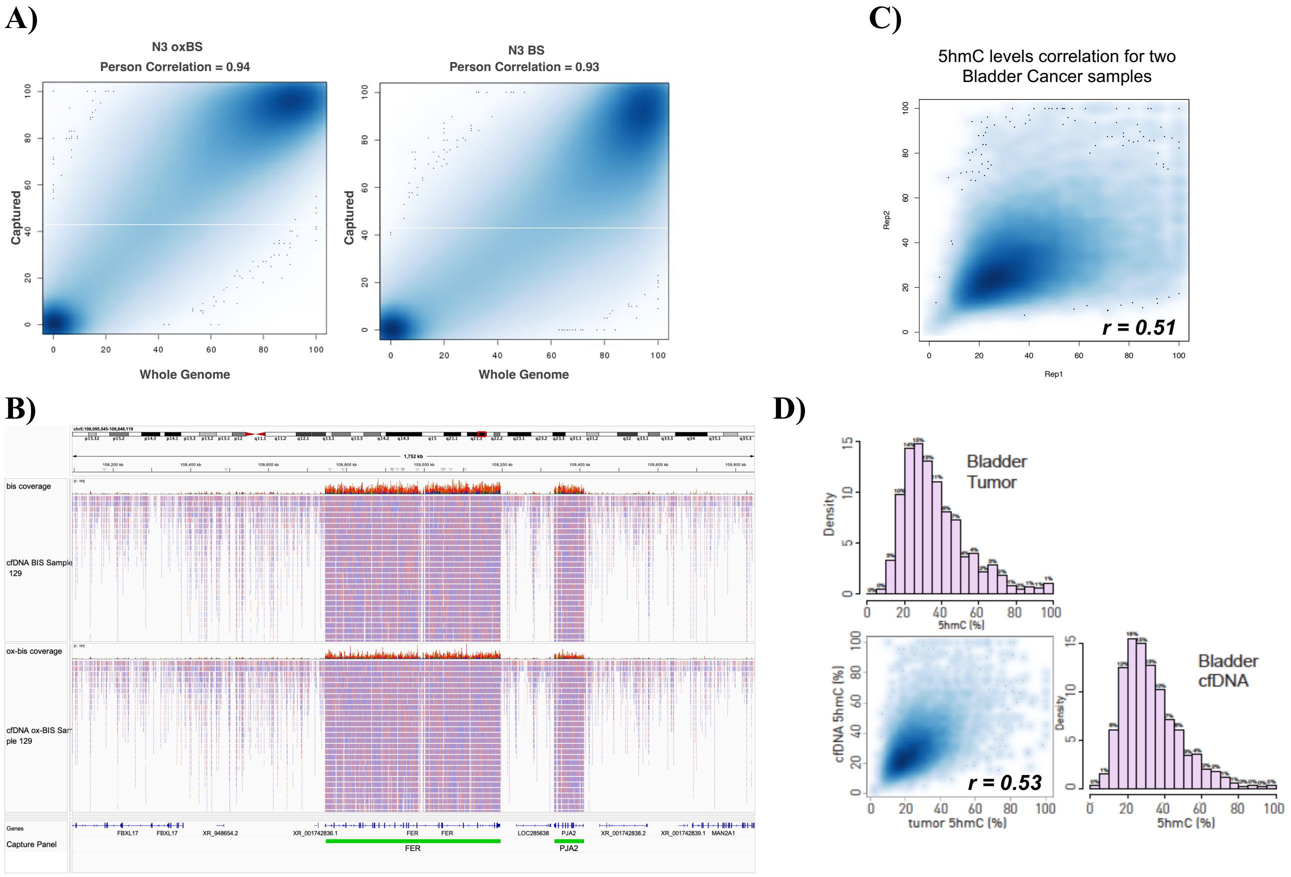
